# Computationally efficient meta-analysis of gene-based tests using summary statistics in large-scale genetic studies

**DOI:** 10.1101/2024.12.06.24318617

**Authors:** Tyler Joseph, Joelle Mbatchou, Arkopravo Ghosh, Anthony Marcketta, Christopher Gillies, Jing Tang, Priyanka Nakka, Xinyuan Zhang, Jack Kosmicki, Carlo Sidore, Lauren Gurski, Regeneron Genetics Center, Maya Ghoussaini, Manuel A.R. Ferreira, Gonçalo Abecasis, Jonathan Marchini

## Abstract

Meta-analysis of gene-based tests using single variant summary statistics is a powerful strategy for associating genes with disease. However, current approaches require sharing the covariance matrix between variants for each study and trait of interest. For large-scale studies with many phenotypes, these matrices can be cumbersome to calculate, store, and share. To address this challenge, we present REMETA, an efficient tool for meta-analysis of gene-based tests. REMETA uses a single sparse covariance reference file per study that is rescaled for each phenotype using single variant summary statistics. We develop methods to apply REMETA to binary traits with case-control imbalance, and estimate allele frequencies, genotype counts and effect sizes of burden tests. We demonstrate the performance and advantages of our approach via meta-analysis of 5 traits in 469,376 samples in UK Biobank. The open-source REMETA software tools and framework will facilitate meta-analysis across large scale exome sequencing studies from diverse studies that cannot easily be brought together.

## Introduction

Over the past 10 years large-scale exome-wide association studies (ExWAS) have proven effective at finding genes associated with disease^1^. By focusing on protein-altering variants, ExWAS often provide interpretable association signals that can help identify therapeutic targets and guide the treatment of disease. For example, the discovery of rare protein-coding variants in GPR75 associated with lower BMI suggest inhibiting GPR75 as a potential therapeutic strategy for obesity^2^. Similarly, rare protein-coding variants in CIDEB are associated with protection against liver disease suggesting CIDEB as a therapeutic target^3^. When combined with array genotyping followed by imputation, ExWAS has comparable association power to whole-genome sequencing for single-variant and gene-based tests^4^.

As most protein altering variants are rare, ExWAS try to improve power by combining single variants in a gene into gene-based tests. Different tests will be better powered to detect an association depending on the genetic architecture of a trait. ‘Burden tests’ are widely used for this purpose and have good power when causal variants alter gene function in the same effect direction^5,6^. Variance component tests model the distribution of effect sizes and can have more power when causal variants act in different directions^7^. Alternatively, methods that combine single variant tests into a single p-value can be particularly powerful when there are only a small number of causal variants^8^. Since the true genetic architecture is unknown, several types of gene-based tests can be combined into a single omnibus test to correct for multiple testing^9,10^. A key component of this approach is the use of variant annotations to group protein damaging variants in each gene to test. Predicting the effect of a variant on protein structure is an active area of research, and a variety of annotation resources exist^11–13^.

Meta-analysis of ExWAS across datasets of diverse ancestry increases power for novel drug target discovery. Effect size meta-analysis (ESMA), also known as inverse-variance weighted meta-analysis, can be used to combine burden tests across studies using estimates of effect sizes and their standard errors. For other tests that do not produce effect size estimates, such as variance component methods, a variety of p-value meta-analysis (PVMA) methods are available for combining across studies. These approaches, which we refer to as standard meta-analysis, are simple and quick to apply, and produce reasonable results^2,3^.

However, meta-analysis of gene-based tests can be challenging if the contributing studies use different annotation resources or different criteria to group variants for testing. Inconsistencies in the variants included in tests can complicate downstream interpretation and analysis. For example, many tests use an allele frequency threshold to include a variant in a gene set. Differences in allele frequencies across studies can change the variants selected in each study. In addition, annotation resources evolve as new approaches are developed. Updating a meta-analysis to use new annotations requires re-analysis of all genes across all studies and traits, which can be significantly costly and time consuming.

These problems can be alleviated by using gene-based tests that leverage single variant (SV) summary statistics. Some gene-based tests (like the weighted sum burden test and variance component tests) can be calculated from appropriately scaled effect size estimates (i.e. score statistics) of individual single variants in a gene, and measures of linkage disequilibrium (LD), or covariance, between those SVs^14,15^. Summary statistics can be combined across studies for meta-analysis, allowing for fine-scale control of the variants included in a test without any repeated association analysis (i.e. no need to go back to the raw genetic or phenotypic data). Gene-based testing from summary statistics can be used to test each gene marginally (or unconditionally), or conditional upon a set of specified variants. The software programs RAREMETAL^15^ and metaSTAAR software^16^ implement this approach. However the LD information required varies according to the exact set of individuals and trait being analyzed. Thus, an LD-like matrix must be computed for each study and trait---which can be challenging to calculate, store and manipulate for studies with large numbers of traits.

In this paper we describe a new approach to this problem that has several key properties.

First, we show that reference LD files derived from all individuals in a study can be accurately substituted for the exact LD files for a given study, even if it includes just a subset of individuals. Such reference LD matrices can be pre-calculated once for a study and used for subsequent analyses which can substantially reduce the compute and storage requirements of gene-based tests. This also greatly facilitates sharing of LD files between groups of researchers since only one LD file needs be shared per study, and not per phenotype and study. We also show that the LD files can be stored sparsely by ignoring LD values between pairs of variants that are close to zero without any loss in accuracy. When all LD is ignored between variants the tests remain relatively accurate at many genes, but we observe some inflation in p-values. Gene-based tests that condition on nearby common variants can help identify which associations are shadows of common variant signals, and in this scenario using LD is crucial.

Second, we have developed a compact per-chromosome binary file format for efficiently storing and sharing per study LD matrices that are required for gene-based tests. This format handles both marginal and conditional testing scenarios and is indexed to allow fast access to the LD information of any gene.

Third, since p-values are not sufficient for follow up interpretation of gene-based tests, we develop an approximate method for calculating allele frequencies, genotype counts and effect size of burden tests from summary statistics.

Fourth, we extend the approach to handle binary trait meta-analysis of gene-based tests with high case-control imbalance and show that this is well calibrated.

Finally, for ease of use we have developed this approach in an open-source software package called REMETA (**URLs**), that is designed to integrate seamlessly with the summary statistic output files from the widely used REGENIE software. REMETA also includes functionality to carry out meta-analysis of gene-based tests using ESMA and PVMA that does not require LD files for each study.

## Results

### Overview of methods

The REGENIE/REMETA workflow is applicable to the setting where there are *P* phenotypes measured in *S* studies with array genotypes and WES data at *K* genes, and proceeds via the following three steps (**Figure 1**).

**Figure 1.**
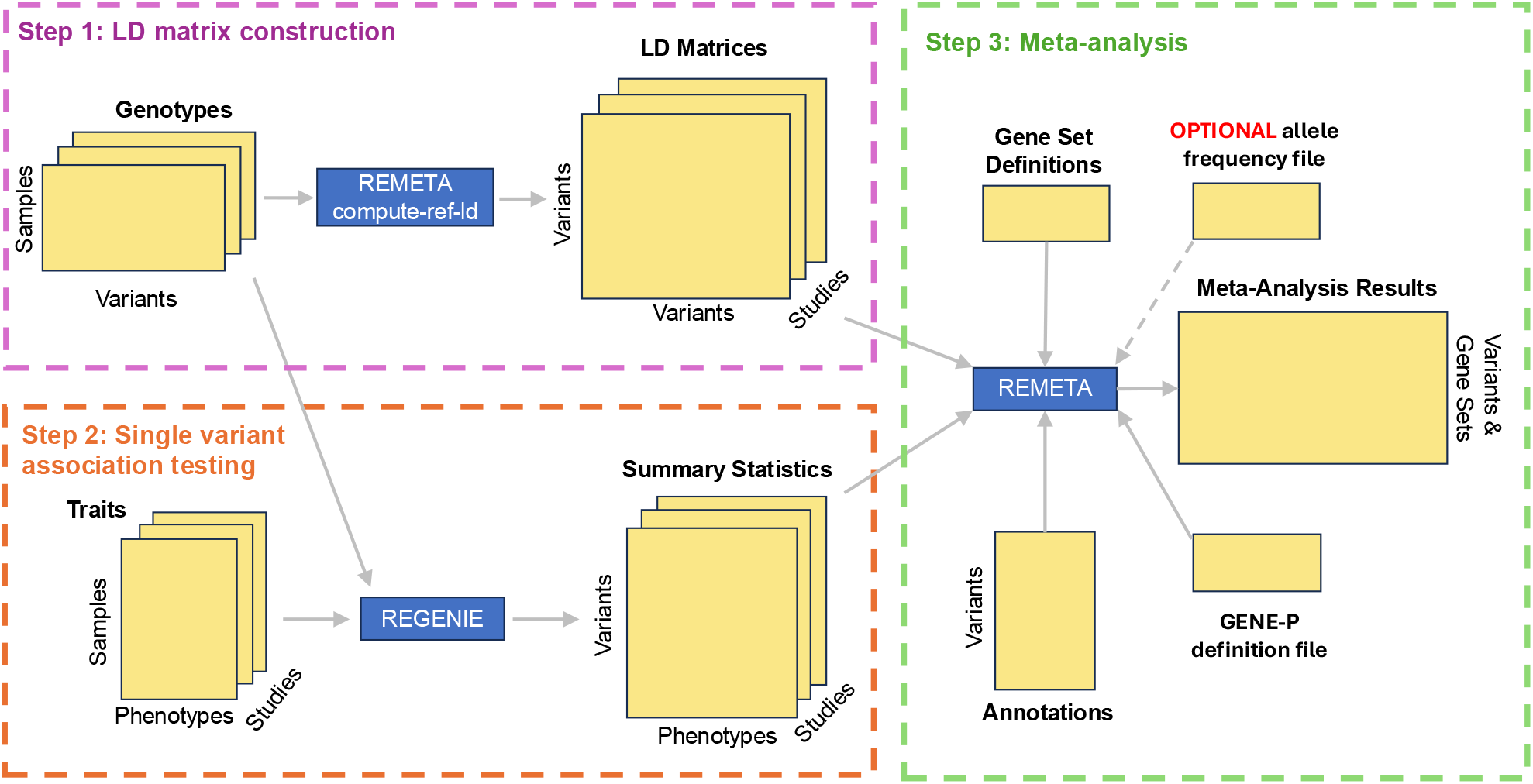
Overview of REMETA’s workflow. REMETA’s workflow has three steps: 1) single variant association testing with REGENIE, 2) an LD matrix construction in REMETA, and 3) meta-analysis.

#### LD matrix construction

The first step constructs reference LD matrices for each study in REMETA. This step only needs to be carried out once for each study. Separate matrices for each of the *P* phenotypes are not required, as with existing approaches^16^, which substantially reduces compute cost and storage. We developed a custom file binary file format to store the LD. The files are indexed so that LD matrices for individual genes can be quickly extracted. The LD files can be constructed from just the WES dataset, which is sufficient for testing each gene marginally. For gene-based analysis conditional upon a set of GWAS loci, the LD files can be constructed from the WES dataset and a file of imputed variants across the genome. Only LD between variants over a threshold value are stored and leads to very compact files with no loss in downstream accuracy.

#### Single variant association testing

The second step involves running REGENIE Step 1 on the array genotypes for each study for the *P* phenotypes with any appropriate covariates. This step accounts for relatedness, population structure and polygenicity. The polygenic scores produced are then used as additional covariates in REGENIE Step 2 where association testing of individual variants in the WES dataset is carried out for each phenotype. It is important to note that *all polymorphic variants* must be analyzed in this step, without any filter on minor allele count. Exclusion of any variants at this step will mean they cannot be included in any downstream gene-based test using REMETA. One key advantage of using REGENIE is that multiple traits can be run in parallel, which controls computational expense and can be simpler to run. We have added the flag --htp flag to Step 2 of REGENIE to produce more detailed summary statistic output required by REMETA.

#### Meta-analysis

The third step carries out gene-based meta-analysis using REMETA (see **Methods**). The inputs to this step are the REGENIE summary statistic files for each trait and study, the REMETA LD files for each study, gene set and variant annotation files, and an optional list of variants to condition on. The format of the gene set and variant annotation files is the same as used by REGENIE and will be familiar to most users. Analysis can be run at all *K* genes, or a smaller specified list of genes, which can be useful when examining associations of interest. REMETA uses the reference LD files for each study and applies a scaling and transformation that depends upon the number of samples included in association analysis of a particular trait (see **Methods**). Burden tests (using the Weighted Sum Test), SKAT variance component tests, and ACATV tests can all be calculated, and also combined into an overall “GENE P” p-value for each gene. The tests can be run on a set of allele frequency bins specified by the user. Overall, this step provides users with fine-scale control of how meta-analysis is carried out.

### Approximate gene-based testing using per study LD

The gene-based tests we consider are constructed from the score statistics of single variants. For burden testing and SKAT-O, computing a p-value requires the covariance matrix of the score statistics to account for the LD of the variants in the test (see **Supplementary Note**). This matrix varies per trait, which can be cumbersome to compute and store across many traits. We therefore wanted to reduce the storage requirements from one matrix per trait to one matrix per study.

In the special case of an association model that tests a single SNP with just an intercept as a covariate, the covariance matrix of the score statistics is equivalent to a rescaling of the covariance matrix of the genotypes in a test^14,17^. Therefore, a natural strategy is to compute the covariance of the genotypes once per study, then adjust it for the trait being analyzed. To make the adjustment, we store the variance of the score statistics computed per trait (see **Methods**). Intuitively, the adjustment corrects for differences in sample size and phenotypic variance across traits. For a gene-based test comprising *p* variants across *T* traits, this reduces the storage requirements from 𝒪(*Tp*^2^+*Tp*) (one *p*×*p* matrix per trait plus the score statistics for each variant) to 𝒪(*p*^2^+2*Tp*) (one *p*×*p* matrix per study plus the score statistics and their variance). To further reduce the storage requirements the covariance matrix can be stored sparsely, keeping only entries between pairs of exome variants where *r*^2^ > 10^−4^ (adjustable through a command line parameter). We refer to the sparse covariance of the genotypes computed once per study as the “reference LD” matrix, and the covariance of the score statistics as the “exact LD” matrix. The goal is to assess how well *p*-values computed using the reference LD matrix approximate those from the exact LD matrix.

We evaluated the performance of this approximation across a range of scenarios using 5 traits in the UK Biobank (n=469,376): BMI, LDL, Breast Cancer (case-control ratio 1:25), Colorectal Cancer (case-control ratio 1:69), and Thyroid Cancer (case-control ratio 1:630). Variants were categorized into 7 annotation groups for gene-based tests: predicted loss of function (pLoF), deleterious missense, possibly deleterious missense, all missense, and combinations of pLoFs with each missense category (**Methods**). Additionally, variants were divided into 5 allele frequency bins (AAF <1%, <0.1%, <0.001%, and singletons). Burden tests were computed across all allele frequency bins resulting in 35 combinations, while SKATO and ACATV were computed for the 1% frequency bin resulting in 7 combinations each.

We found that approximate *p*-values are accurate across a wide range of settings. **Supplementary Figure 1 (top row)** shows the performance of the approximation when the reference LD is calculated in the full UKB samples (ALL N=469,376) and used to calculate burden test *p*-values (specifically the sum test^5^: see **Methods**) for BMI (N=467,484), LDL (N=446,939), Breast Cancer (N=436,422), Colorectal Cancer (N=437,212) and Thyroid Cancer (N=437,417). As the LD can vary among genetic ancestries, we repeated this experiment in subsets of UK biobank with different ancestries: EUR (N=445,418), AFR (N=9,295) and SAS (N=10,614). In each case we computed reference LD matrices within each ancestry, and compared *p*-values computed with reference LD to those computed using the exact LD matrix (**Supplementary Figure** 1; remaining rows). All these scenarios have the property that the sample sizes for the traits being analyzed are close to the size of the full reference panel, and show excellent agreement. **Supplementary Figure 2** shows analogous results for the SKAT-O test (see **Methods**).

The approximation can also be used when the sample of trait being analyzed is a smaller subset of the full reference panel. We hypothesized that the approximation might break down when the subset was very small and/or the ancestry composition of the subset was different that of the full reference panel. **Supplementary Figure 3 (top row)** shows the performance of the approximation when the reference LD is calculated in the full UKB samples (ALL N=469,376) and used to calculate sum test p-values in a random subset about 1/3^rd^ of the size : BMI (N=155,832), LDL (N=148,977), Breast Cancer (N=145,628), Colorectal Cancer (N=145,866) and Thyroid Cancer (N=145,938). The other rows show the accuracy when analyzing different ancestry subsets. For example, the third and fourth rows show results for the rather extreme scenarios where the small subsets of SAS (mean N=10,017) and AFR (mean N=8,602) samples are analyzed using reference LD calculated from ALL UKB samples. In all cases the agreement is very good. **Supplementary Figure 4** shows analogous results for the SKATO test.

One possible explanation for the strong concordance between *p*-values is that there is little or no LD between the exome variants in a test. In the absence of LD between variants, the reference LD matrix is equal to the exact LD matrix, and there would be no need to store LD matrices per study. Therefore, we compared *p*-values computed using the exact LD matrix to *p*-values computed by ignoring any LD between variants (**Supplementary Figure 5**). When LD between variants is ignored, tests statistics at many genes are unaffected, but we observed inflation in the *p*-values of some tests at some genes, suggesting that LD between variants is required to have well-calibrated tests. When carrying out conditional tests of association using LD is crucial.

### Estimating genotype counts, allele frequencies, and effect sizes

For burden testing REMETA, our goal is to approximate meta-analysis of burden tests from REGENIE. We therefore sought to estimate genotype counts, allele frequencies, and effect sizes of REGENIE’s burden masks. The default burden test in REGENIE is performed using the collapsing variant test. In contrast, REMETA uses the sum test^5,18^ to construct burden masks, as it can be computed from single variant summary statistics. Despite differences in how the tests are computed, the *p*-values from both tests are concordant (**Supplementary Figures 6-7)**, motivating us to use the sum test as an approximation to the collapsing variant test. We derived new methods to estimate genotype counts and allele frequencies of the burden masks for collapsing variant test from the single variant frequencies and reference LD (see **Methods**) and show that our estimator accurately approximates allele frequencies and genotype counts computed in REGENIE (**Supplementary Figures 8-10**). Genotype counts were particularly accurate for burden masks comprising pLoF variants (**Supplementary Figures 11-12**). We compared our method of estimating genotype counts to a naïve estimate that sums the genotype counts of the mask variants. As expected, the naïve estimator overestimates burden mask genotype counts (**Supplementary Figure 13**). Finally, we derived estimates of the effect size for the sum test (see **Methods**), and show that it accurately approximates the collapsing variant test estimate as the standard errors of both estimators decrease (**Supplementary Figure 14**).

### Computational and storage cost of REMETA

Reassured that gene-based tests can be approximated from sparse per study LD matrices, we developed a compact file format to store LD matrices. LD matrices can be computed for either marginal testing by only including exome variants, or for conditional analysis by including exome variants and imputed variants in a buffer region around each gene (**Figure 2A**). Because generating LD matrices between large numbers of variants can be compute and storage intensive, we explored several options to improve scalability. To speed up compute time, REMETA performs matrix multiplication using single-precision floating-point numbers instead of double-precision floating point numbers. Using Advanced Vector Extensions (i.e. AVX) for x86 architectures, 8 single-precision floating-point numbers can be multiplied simultaneously, doubling the speed of matrix multiplication compared to 4 double-precision floating-point numbers. To reduce LD file sizes, REMETA supports storing 3 floating point types: 4-byte single precision, 2-byte and 1-byte floating-point numbers (**Figure 2B**). The output LD matrices are compressed using HTSlib with the DEFLATE library which provides heavily optimized gzip compression and indexed per gene to quickly query LD matrices per gene.

**Figure 2.**
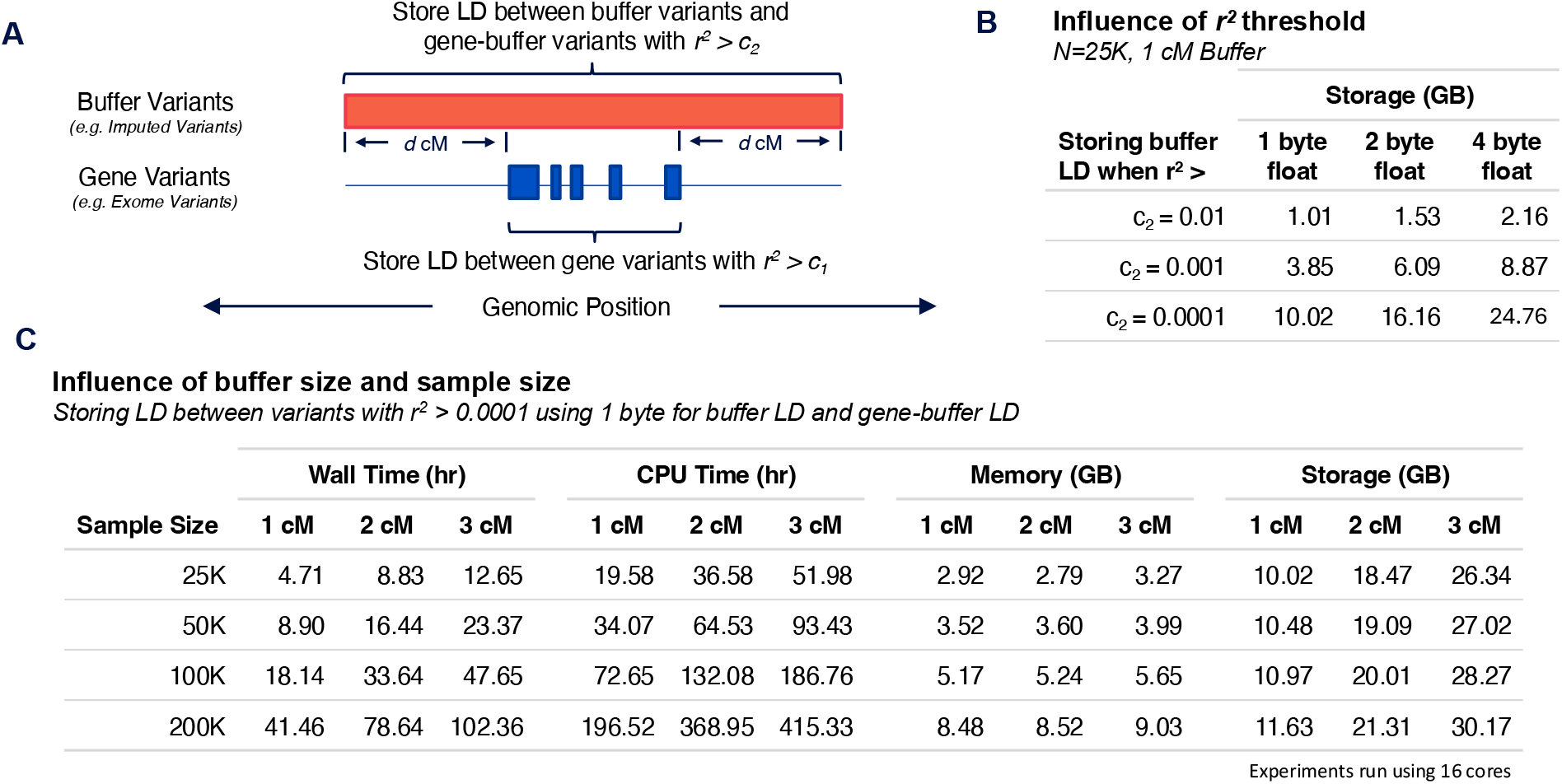
Resource requirements to generate LD matrices for conditional analysis. Experiments were performed using chromosome 20 in UK Biobank. **(A)** Depiction of REMETA’s strategy for computing LD matrices. **(B)** How the r^2^ threshold influences LD file sizes for three floating point storage types. **(C)** Wall time, CPU time, memory usage, and storage requirements to generate LD matrices in subsets of UK Biobank.

**Figure 2C** illustrates the scalability of LD matrix generation in REMETA with varying sample and buffer sizes in subsets of the UK Biobank. REMETA’s main constraint is compute time, which approximately doubles for each doubling in sample size and increases linearly with buffer size. In contrast, memory and storage requirements increase more slowly with sample size. Across all experiments REMETA used <9GB of memory. LD file sizes were comparable across sample sizes, reflecting the similar numbers of imputed variants in each subset.

We benchmarked LD matrix generation for marginal analysis against MetaSTAAR by computing LD matrices in subsets of UK Biobank (**Table 1**). Because MetaSTAAR computes LD matrices per trait, at each sample size we simulated a null quantitative trait from a standard normal distribution without covariates. REMETA, in contrast, computes a single matrix per study that is reused across traits. We compared CPU time, memory usage, and file storage between REMETA and MetaSTAAR. At 200K samples, we found that REMETA was >2.5x faster than MetaSTAAR, used 77% less memory, and generated files that were 56% smaller than those generated by MetaSTAAR. Overall, LD matrices for marginal analysis are small and quick to compute because they only require the LD between exome variants in a gene.

**Table 1.** Per gene LD matrices were computed on chromosome 20 using exome variants in subsets of UK Biobank. Note that MetaSTAAR requires an LD matrix per trait in each study while REMETA only requires a single LD matrix per study.

| Sample Size | CPU Time (min) |  | Memory (GB) |  | Storage (MB) |  |
| --- | --- | --- | --- | --- | --- | --- |
|  | REMETA | MetaSTAAR | REMETA | MetaSTAAR | REMETA | MetaSTAAR |
| 25K | 0.95 | 23.55 | 1.34 | 2.77 | 1.16 | 2.67 |
| 50K | 2.93 | 29.81 | 2.26 | 5.41 | 1.41 | 3.10 |
| 100K | 9.15 | 41.15 | 3.95 | 12.33 | 1.73 | 3.86 |
| 200K | 28.06 | 75.16 | 7.17 | 30.63 | 2.16 | 4.93 |

Experiments run using a single core

For conditional analysis, the comparison between MetaSTAAR and REMETA is not so straight forward. MetaSTAAR requires a user to specify a set of variants to condition on *before* the calculation of the LD matrices. As the set of conditioning variants is usually small this does not add appreciably to the compute or file sizes, but it does require a new set of LD matrices every time an analysis changes. In REMETA, we aim to calculate a reference set of LD matrices once, so that the set of variants can be specified and changed at will *after* the calculation of the LD matrices. Nonetheless, REMETA also provides an option to prespecify variants for conditional analysis.

### Conditional analysis

Conditioning gene-based tests on nearby common variants can help identify which associations are shadows of common variant signals due to LD. In conditional analysis from summary statistics, the variants available to condition on depends on the size of the buffer region around each gene and the r^2^ threshold to store elements in the LD matrix. Both parameters influence the size of the matrices stored. We therefore asked: how large do LD matrices need to be to effectively condition out most common variant signals?

We re-analyzed all 157 ExWAS significant 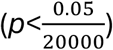 GENE_P^19^ (**Methods**) associations found among the 5 traits in UK Biobank. We computed gene-based tests conditional on index SNPs computed by LD clumping common variants for each trait (MAF>0.01, r^2^>0.1), and compared p-values from REMETA to p-values from REGENIE. Gene-based tests were conditioned on 1,961 index SNPs in BMI, 2,698 index SNPs in LDL, 61 index SNPs in Breast Cancer, 10 index SNPs in Colorectal Cancer, and 2 index SNPs in Thyroid Cancer.

For all traits except LDL REMETA’s conditional p-values were similar across buffer sizes and r^2^ thresholds (**Figure 3 and Supplementary Figure 15**). Conditional analysis for LDL, which had the most index SNPs and some very strong common variant associations, was sensitive to both. At r^2^>0.01, increasing the buffer size increased the number of significant associations in LDL compared with REGENIE (**Figure 3A**), suggesting a more stringent r^2^ threshold is required to condition out common variant signals.

**Figure 3.**
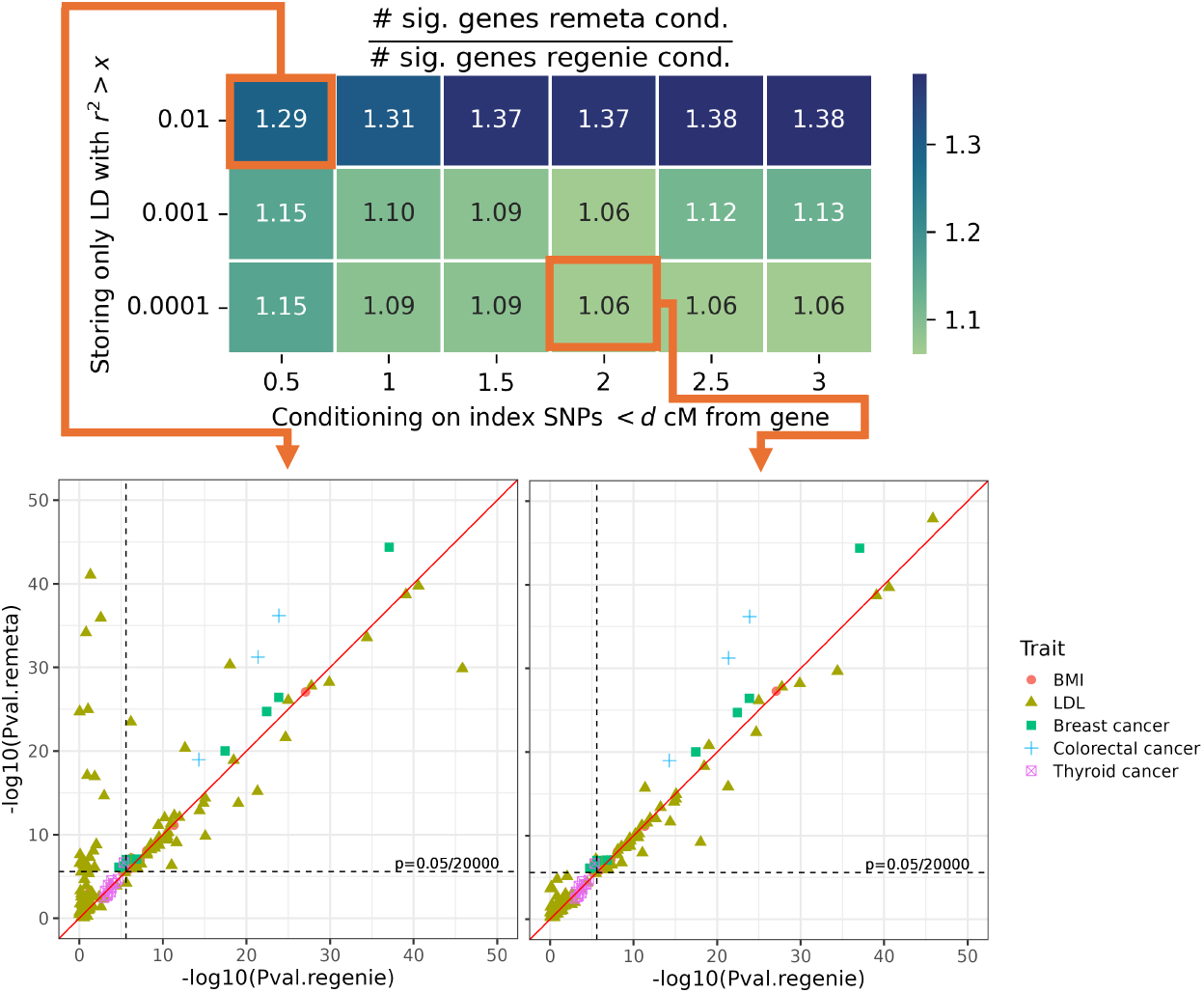
Evaluation of LD buffer size and r^2^ threshold required to condition out common variant signals. Number of significant gene-trait pairs in REMETA compared to REGENIE. REMETA conditional analysis is performed from summary statistics while varying the buffer size and r^2^ threshold. REGENIE conditional analysis is performed by including all index SNPs on a chromosome as covariates. At d=2 and r^2^>0.0001 REMETA conditions out most common variant signals. The remaining significant gene-trait pairs are near the significance cutoff.

Our results suggest that for many traits, including highly polygenic traits like BMI, an r^2^>0.01 and buffer size of 0.5 cM is sufficient to condition out common variant signals. Using these parameters, we can estimate the total file size of LD matrices required for conditional analysis. If LD matrices on chromosome 20 represent 2% of the total file size, then the total storage required for LD matrices genome wide is <60 Gb for N=200K samples. For traits like LDL, an r^2^>0.0001 and cM=2 is required---totaling approximately 1Tb of storage for N=200K samples. The right balance of parameters will depend on the traits to be meta-analyzed.

### Type 1 error and unbalanced binary traits

For unbalanced binary traits using a normal approximation to the score test statistic can lead to inflated type 1 error. In these instances, a saddlepoint approximation (SPA) has proven an effective strategy to help calibrate *p*-values. We use two observations to calibrate *p*-values for unbalanced binary traits. First, the sum test in REMETA approximates the collapsing variant test REGENIE. Second, we can estimate genotype counts of the collapsing variant test from the variants in the test and their LD. This allows us to apply a summary statistics-based SPA^20^ for the collapsing variant test and use it to compute a calibration factor to re-scale the LD matrix (**Methods**).

We evaluated type 1 error for burden tests and SKAT-O in REMETA by simulating null traits for 3 equally sized subsets of UK Biobank. Ten simulation replicates were generated for each of 5 traits: a quantitative trait, and binary traits with case-control ratios ranging 1:1, 1:9, 1:49 and 1:99. Our simulations show that REMETA provides good type 1 error control across traits (**Supplementary Table 2**). For binary traits with case-control ratios 1:1 and 1:9, type 1 error was similar between SPA corrected *p*-values and raw *p*-values. For the more unbalanced binary traits, SPA corrected *p*-values had better type 1 error control than uncorrected *p*-values. SPA corrected *p*-values were well-calibrated for burden testing, and showed improved type 1 error control for SKAT-O.

### Application to meta-analysis of the UK Biobank

We applied REMETA to carry out gene-based meta-analysis of 3 equally sized subsets of UK Biobank with 156K samples each (N=469,376 total). We analyzed two quantitative traits, body mass index (BMI) and low-density lipoprotein (LDL), and three binary traits: Breast Cancer (case-control ratio 1:25), Colon Cancer (case-control ratio 1:69), and Thyroid Cancer (case-control ratio 1:630). For each subset, we generated LD matrices with REMETA and single variant summary statistics with REGENIE.

We compared the performance of REMETA to the standard meta-analysis approach that combines effect sizes for burden tests (ESMA) and combines *p*-values (PVMA) for variance component and ACATV tests. **Supplementary Figures 16-18** show the strong similarity between QQ plots and genomic control measures for the different tests using both meta-analysis approaches. The overall omnibus GENE_P *p*-values^19^ for both models are largely consistent (**Figure 4 top row**), with REMETA identifying more significant gene-trait pairs overall. Across the 5 traits, REMETA identified 119 significant gene-trait associations (at a Bonferroni corrected threshold 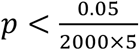) compared to 98 significant gene-trait associations identified by standard meta-analysis (**Figure 5A**). Notably, only 1 significant gene-trait pair in standard meta-analysis was not significant in REMETA (IGSF23 with LDL; standard meta-analysis GENE_P=4.4e-7, REMETA GENE_P=6.2e-7). To better understand which tests contributed to the 22 additional gene-trait pairs identified by REMETA, we counted the most significant test for each pair (**Figure 5B**). SKATO-ACAT was the most significant test for 14 gene-trait pairs, followed by ACATV-ACAT for 8 pairs and BURDEN-ACAT for 2 pairs, suggesting that SKATO and ACATV meta-analysis from summary statistics is more powerful than SKATO and ACATV meta-analysis using PVMA.

**Figure 4.**
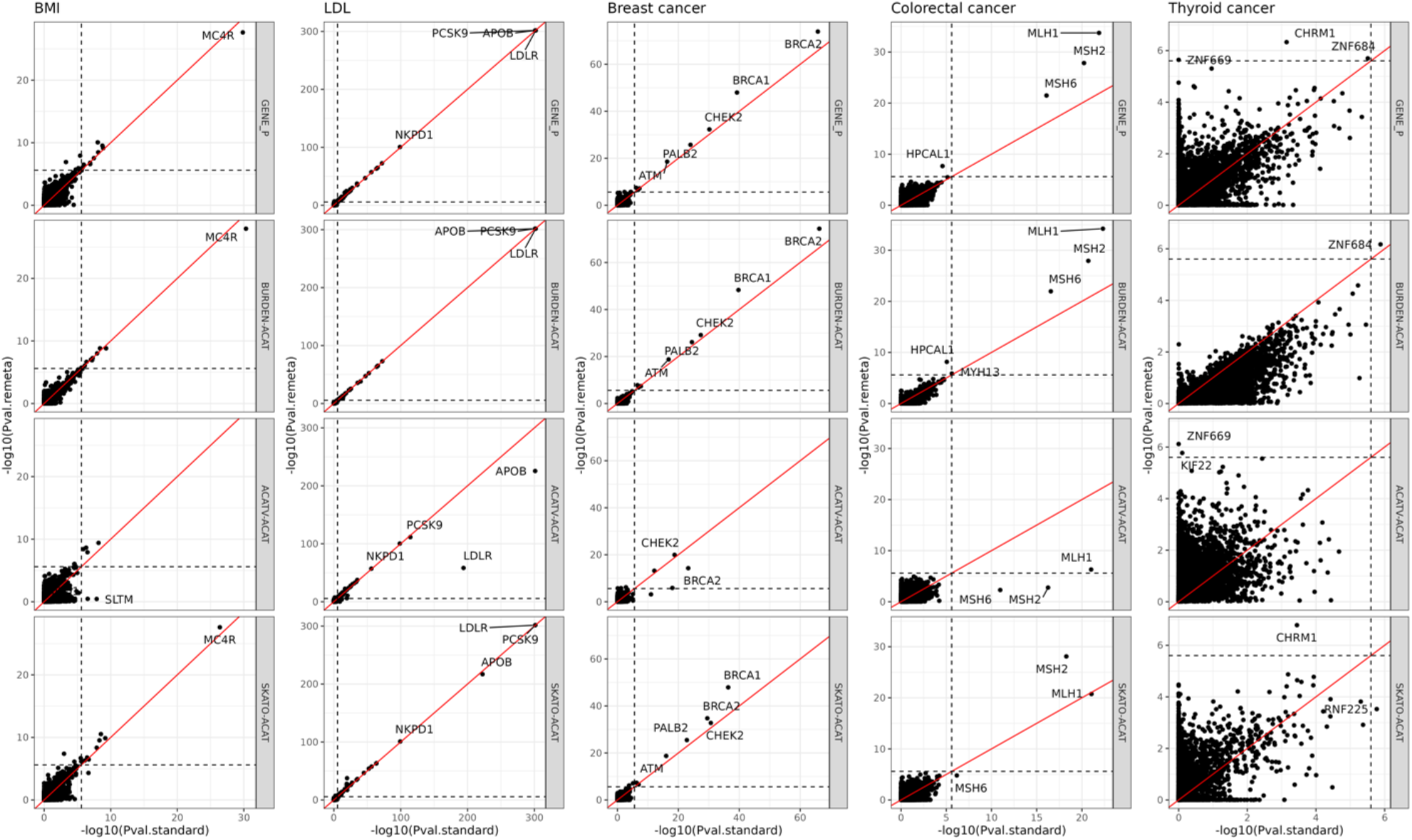
Comparison of –log10 p-values between REMETA to standard meta-analysis across 5 traits. Each point is a single gene, where multiple *p*-values per gene have been combined using ACAT. Four types of gene-based tests are compared: GENE_P, burden, ACATV, and SKATO. \**GENE_P*: Omnibus test combining *p*-values from BURDEN-ACAT, ACATV-ACAT, and SKATO-ACAT using ACAT. \**BURDEN-ACAT*: ACAT of burden meta-analysis *p*-values. \**ACATV-ACAT*: ACAT of ACATV meta-analysis *p*-values per gene. SKATO-ACAT: ACAT of SKATO meta-analysis *p*-values per gene. Dashed lines correspond to a *p*-value of 2.5×10^-6^.

**Figure 5.**
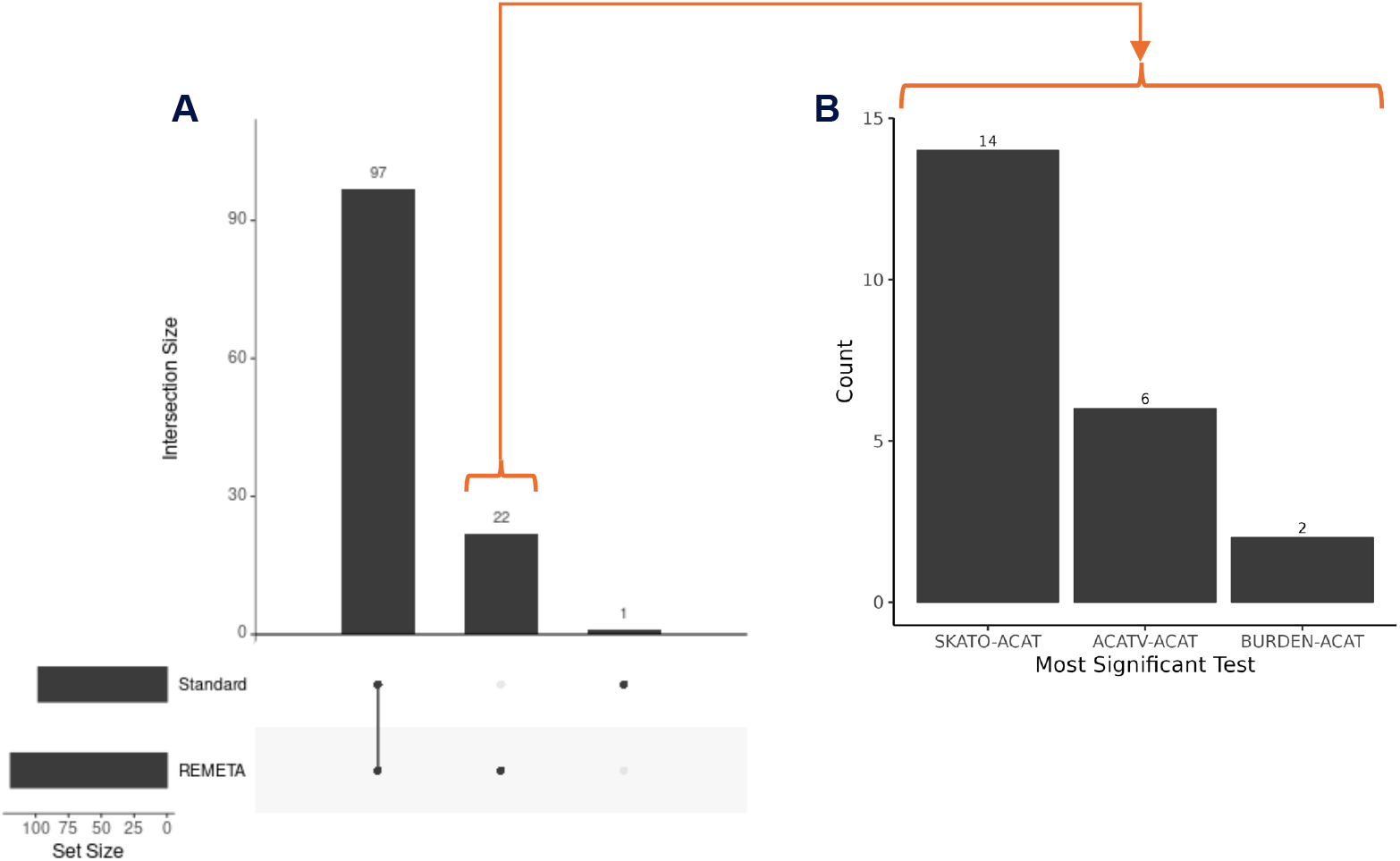
Significant gene-trait associations in REMETA and standard meta-analysis. **(A)** Number of GENE_P significant gene-trait associations 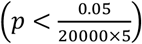 from REMETA and standard meta-analysis across 5 traits in UK Biobank. **(B)** Most significant test included in GENE_P for each of 22 gene-trait pairs that were significant in REMETA alone.

For ACATV, we observed associations stronger associations in standard meta-analysis than REMETA between LDLR in BMI, and ATM, BRCA1 and BRCA2 in Breast Cancer, and MSH2, MSH6 and MLH1 in Colorectal Cancer (**Figure 4 third row**). These associations are driven by collapsing low minor allele count variants into a single meta-variant and including them in ACAT-V. However, signals driven by variant collapsing are similar to burden signals, so these genes get picked by other tests. This suggests that variant collapsing in ACAT-V does not add much power when combined with other gene-based tests.

For burden tests, we observed stronger associations in REMETA than standard meta-analysis between BRCA1 and BRCA2 in Breast Cancer, and MLH1, MSH2, and MSH6 in Colorectal cancer (**Figure 4 second row**). These are likely due to differences in testing for association from summary statistics compared to individual data for binary traits. In REGENIE, variants are first collapsed, then tested for association using Firth regression. In REMETA, single variant summary statistics are computed using Firth regression, then tested for association by combining summary statistics. This can lead to differences in p-values between the two methods (**Supplementary Figures 19-20**), though QQ plots for both methods remain similar (**Supplementary Figures 21**).

### Fast re-analysis using new variant annotations

To illustrate the fast and flexible properties of the REMETA approach, we re-analyzed gene-based tests using ESM1v protein language model annotations of missense variants^13^ (**Methods**). As variant annotation approaches continue to evolve repeated re-analysis of datasets is likely, thus it is desirable to make this as simple as possible. We benchmarked how long re-analysis would take in REGENIE using the raw genotype and phenotype data, compared to REMETA using the summary statistics and LD for chromosome 20 in BMI. Re-analysis took 162.9 CPU minutes (∼54.3 CPU minutes per batch) in REGENIE and 7.94 CPU minutes in REMETA. Encouraged by the computational performance of REMETA, we re-analyzed all 5 traits using ESM1v missense annotations. We highlight three gene-trait associations with the largest differences in association strength between GENE_P p-values computed with and without the additional ESM1v masks: MC4R with BMI (GENE_P without ESM1v = 2.3×10^-28^, GENE_P with ESM1v = 3.8×10^-38^), ANGPTL3 with LDL (GENE_P without ESM1v = 4.0×10^-73^, GENE_P with ESM1v = 6.7×10^-80^), and BRCA1 with Breast Cancer (GENE_P without ESM1v =1.1×10^-48^, GENE_P with ESM1v = 9.3×10^-56^). All are well-known associations with improved significance when including ESM1v masks.

## Discussion

In this study, we proposed REMETA as a computationally efficient approach to meta-analysis of gene-based tests in exome-wide association studies. The software integrates seamlessly with the widely used REGENIE software for carrying out individual study ExWAS. Two aspects of REMETA allow it to scale to multiple large studies like UK Biobank. First, REMETA uses a single LD matrix computed per study instead of one LD matrix per trait. We showed under a range of scenarios that one LD matrix per study is sufficient to perform gene-based testing from summary statistics. Second, REMETA efficiently computes and stores per study LD matrices for each gene. To aid in the interpretation of gene-based association results, we developed methods to estimate the allele frequencies, genotype counts, and effect sizes of burden tests from single variant summary statistics, whereas existing approaches have only been able to provide p-values.

REMETA allows full flexibility to define annotations at meta-analysis without the need to re-analyze individual level data. There are several benefits to this approach. As annotation resources continue to evolve, the ability to easily update meta-analysis results will aid in the discovery of new associations. For example, we showed that better missense predictions from the ESM1v protein language model led to more significant associations between MC4R with BMI, ANGPLT3 with LDL, and BRCA1 with Breast Cancer. These examples highlight the importance of the ability to re-analyze gene-based tests with new annotation resources. Similarly, having carried out a meta-analysis of several studies using REMETA, adding in a new study is then very easy. Furthermore, we showed re-analysis of gene-based tests with REMETA is considerably faster than re-analysis of individual level data. Thus, REMETA is well-suited to re-analysis across large numbers of traits like those found in biobanks or large health systems.

There are also limitations to the REMETA approach. For example, any update to the phenotypes, or changes to the covariates, requires recomputing single variant summary statistics from the original data in REGENIE. It may be possible to incorporate new covariates through adjusting the reference LD matrix, though we leave this for future work. Additionally, while we showed that LD between variants was not critical for marginal testing, accurate LD is important for conditional analysis and estimating genotype counts. Consequently, the size of the LD files for conditional analysis were large compared to those for marginal analysis.

The REMETA software tools and framework will facilitate meta-analysis across biobank datasets that cannot easily be brought together via sharing of appropriate summary statistic files. For example, the apparent move towards biobank datasets being hosted on distinct trusted research environments (TREs) such the UK Biobank RAP and the AllofUs Researcher Workbench (**URLs**) means that these datasets are highly unlikely ever to be combined in a single environment. Centralized calculation of reference LD files for such studies that are then shared freely with researchers would greatly facilitate the ease with which meta-analysis across studies could occur.

## Methods

### Computing gene-based tests from summary statistics

We considered three gene-based tests that can be computed from single variant summary statistics: the weighted sum test (WST), the optimal sequence kernel association test (SKAT-O), and the aggregated Cauchy association test (ACAT-V). For a single gene-based test in a single study, the setup is as follows. For a sample of *n* individuals, let *y* be the *n*×1 vector of phenotypes, *G* the *n*×*p* genotype matrix of variants in the test, and *w* a *p*×1 vector of variant weights.

Both the WST and SKAT-O can be computed from the score statistics^18^ (using linear regression for quantitative traits and logistic regression for binary traits) of the variants included in the test and their covariance

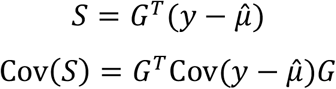

where 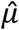 is estimated under the null hypothesis.

The test statistic for WST is

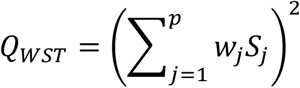

where the *w*_*j*_ are prespecified variant weights. We refer to the special case where *w*_*j*_ = 1 as simply the “sum-test”. Then

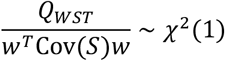

The test statistic for SKAT-O is

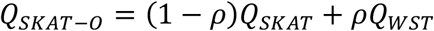

Where

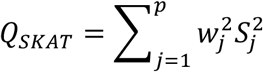

The test statistic *Q*_*SKAT*−0_ follows a mixture of chi-square distribution (see **Supplementary Note**). Similar to the original SKAT-O method, we compute *p*-values for SKAT-O using a series of values for ρ ∈ (0, 0.01, 0.04, 0.09, 0.16, 0.25, 0.5, 1). However, instead of taking the minimum *p-*value across ρ values we combine *p*-values using ACAT.

For ACAT-V, we first compute the marginal association p-values *p*_*j*_ for each variant *j* from the score statistics *S*_*j*_ and their variance. Then the *p*-values are combined using the test statistic

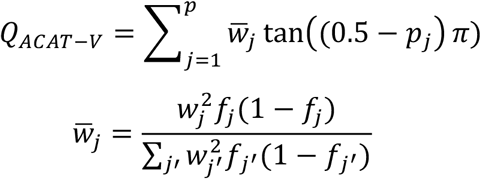

*Q*_*ACAT−V*_ follows a standard Cauchy distribution (see **Supplementary Note**).

REMETA, like MetaSTAAR, combines p-values across gene-based tests into a single omnibus test termed “GENE_ P.” GENE P *p*-values are computed in two steps. In the first step, *p*-values are combined per test using ACAT into BURDEN-ACAT, ACATV-ACAT, and SKATO-ACAT p-values. In the second step per test p-values are combined with ACAT to compute GENE P.

### Approximate gene-based testing using per study LD

The matrix Cov(*S*) needs to be computed and stored for every trait and every study in the analysis. We therefore wanted to approximate Cov(*S*) with a matrix computed once per study. As others have noted^14,17^, in the absence of covariates (except an intercept) Cov(*S*) = νCov(*G*^T^) where ν is a scalar that depends on the phenotype. This suggests we should compute Cov(*G*^*T*^) once, then adjust it for the phenotype being testing. A natural choice is to adjust Cov(*G*^*T*^) to have the same diagonal as Cov(*S*). This is the approach we take.

Let *G* be the *n*×*p* genotype matrix of all variants in a gene, *S*_*t*_ be the score statistics of all variants in a gene for a particular trait *t*, and Φ_*t*_ = Cov(*S*_*t*_) be their covariance. We propose storing three pieces of information to construct gene-based tests: *S*_*t*_, *D* = diag(Φ_*t*_), and Cov(G^*T*^). Let *D* be the diagonal matrix with entries diag(Φ_*t*_). We then approximate (note that Corr(*G*^*T*^) is easy to compute from Cov(*G*^*T*^)):

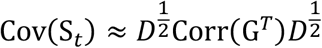

Thus, for *T* traits the time and space requirements are reduced from *G*(*Tp*^2^+*Tp*) (*T* matrices of size *p*×*p* plus the score statistics) to *G*(*p*^2^+2*Tp*) (one matrix of size *p*×*p*, the score statistics, and the diagonal of Φ_*t*_).

### Effect size meta-analysis (ESMA)

Effect sizes are meta-analyzed using an inverse-variance weighted meta-analysis. Specifically, if *β*_1_, …, *β*_*K*_ and *SE*_1_, …, *SE*_*K*_ are the effect sizes and standard errors for a variant across *K* studies, we compute

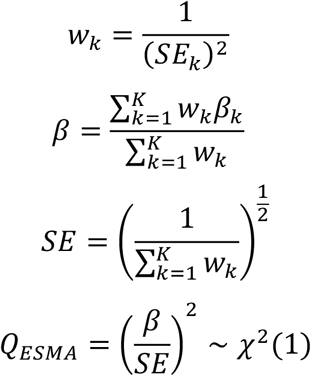

### P-value meta-analysis (PVMA)

P-value meta-analysis is performed using Stouffer’s method. In Stouffer’s method, p-values are first converted to z-scores, then combined by taking a sum weighted by the sample sizes of each study. Let *p*_1_, …, *p*_*k*_ be the p-values for a test across *K* studies with sample sizes *N*_1_, …, *N*_*k*_. We compute

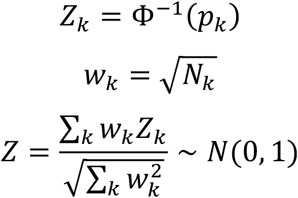

### Meta-analysis of gene-based tests

Let *S*_*k*_ be the *j*×1 vector of score statistics for variants in a gene-based test in study *k*. For meta-analysis of WST and SKAT-O across studies, we can combine the score statistics across studies

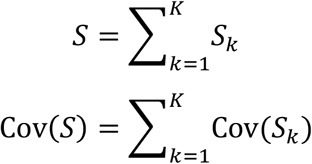

Then *p*-values can be computed like the single study case above.

### Estimation of effect sizes

In the sum test, each variant in a mask is assumed to have the same effect size. Consequently, if *β* is the true effect size to be estimated, and 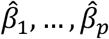 are the marginal effect size estimates of the variants in the mask, then 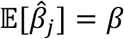 for each variant *j* in the mask. Furthermore, for any convex combination *w*_1_, …, *w*_*p*_ of the effect sizes we have 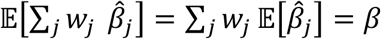. We could use this observation to look for an estimator that minimizes 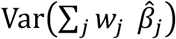. If there is no correlation between the effect sizes in the mask, this is equivalent to an inverse-variance weighted meta-analysis of the variants in the mask. However this would require repeatedly solving a non-negative least squares problem for each burden test, which is computationally expensive. Therefore, we set 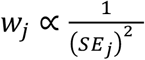, the weights used by inverse-variance meta-analysis. If there is little LD between the variants in the mask, then the inverse-variance weighted estimator and minimum variance estimator should be similar.

For meta-analysis of burden tests, we first estimate effect sizes and standard errors per study. Then we combine effect sizes and standard errors across studies using inverse-variance weighted meta-analysis.

### Conditional analysis

REMETA stores the LD of variants within a gene and in a user specified buffer region around each gene. So long as a variant is stored in the LD matrix of a gene it can be used to perform conditional analysis of gene-based tests. If we let *S*_G_ and *S*_*c*_ be the score statistics in a gene-based test and the score statistics of variants to condition on, then under the null hypothesis

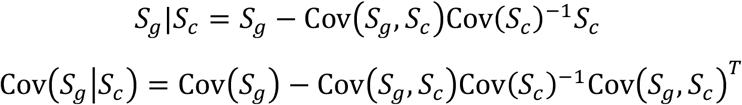

For meta-analysis, we perform conditional analysis across all studies at once ---i.e. on the sum of score statistics across studies.

We also estimate the conditional effect sizes of gene-based tests. If *β_G_* and *β*_*c*_ are the effect size estimates for the variants in a gene-based test and effect size estimates of the variants to condition on, then we compute

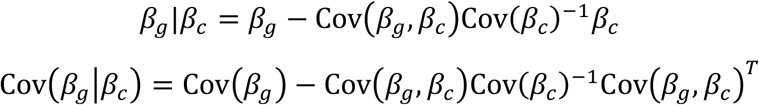

We estimate the covariance of 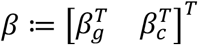 using a similar strategy to the approximation for score statistics. If *D* = diag{Cov(*β*)}, then we approximate 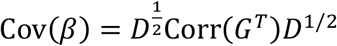 where *G* is the *n*×*p* matrix of variants in the test. This is similar to the strategy used by COJO^21^.

For meta-analysis, we first condition the effect sizes within each study, then combine them using the method described in “Estimation of effect sizes.”

### Estimation of genotype counts and allele frequencies

If genotype counts of the single variants are known, then we can use them to estimate genotype counts of the burden mask. Let *G*_1_, …, *G*_*p*_ be the vectors of genotypes at the *p* variants in the mask, and let *Y* = max{*G*_1_, …, *G*_*p*_} ∈ {0,1,2} be the mask genotype vector calculated by taking the maximum genotype across elements of the variant vectors. Additionally, let 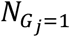 be the number of individuals who are heterozygotes at variant *j*, and *N*_*Y*=1_ be the number of individuals who are heterozygous for the burden mask. We want to estimate *N*_*Y*=1_ from 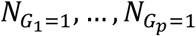. Specifically, we want to find coefficients *c*_*j*_ such that

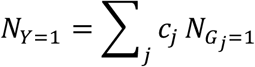

The strategy we take is to compute *c*_*j*_ sequentially, such that each *c*_*j*_ estimates the proportion of the 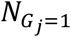 we have yet to count. Specifically,

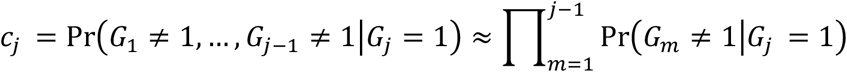

The approximation is chosen because the terms Pr(*G*_*m*_ ≠ 1|*G*_*j*+1_ = 1) can be computed from the LD matrix (the case for homozygotes is similar, see **Supplementary Note**).

### Extension to binary traits with case-control imbalance

For unbalanced binary traits, using a normal approximation to the distribution of the score statistic can lead to inflated type I error. Saddlepoint approximation (SPA) has been shown to be an effective strategy for controlling type I error for both single variants^22^ and gene-based tests ^23^. SPA uses the cumulant generating function of the score statistic to approximate its null distribution. For a variant *j*, the cumulant generating function of the score statistic for logistic regression is

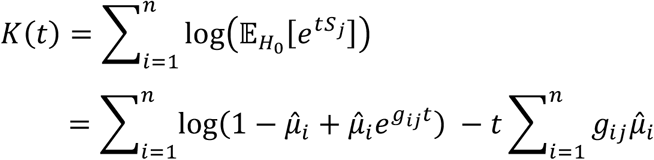

where 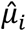 is estimated under the null hypothesis. In the setting of single variant meta-analysis, SPA has been extended a summary statistics approach by recognizing that, if the model only includes an intercept, then an SPA can be computed from case-control counts and genotype counts alone^20^.

In REMETA, we use the observation that the sum test approximates the collapsing variant test, and that we can we estimate genotype counts of for collapsing variant test (see details in **Supplementary Note**). Briefly, for a study *s* let 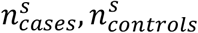 be the number of case and number of controls. Let 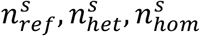 be the number of homozygous reference, heterozygous, and homozygous alternate genotypes for a mask. Then we have

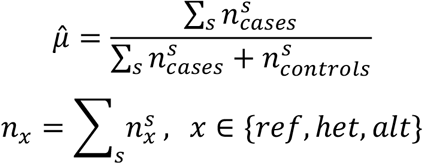

We use these terms to fit an SPA to the collapsing variant test. The cumulant generating function for the SPA is

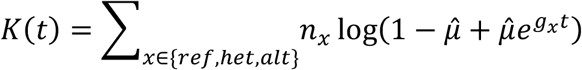

where *g*_*x*_ is the mean centered genotype divided by 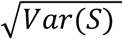. We use this SPA to compute an approximate p-value for the sum test.

For SKATO, we use the SPA computed for the sum test to compute a calibration factor using a similar strategy to^24^. Let *p*_*ST*_ and *p*_*SPA*_ be the p-values computed from the sum test and SPA respectively. Let 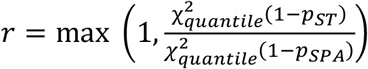, then we replace the matrix Cov(*S*) by *r*Cov(*S*).

### Variant annotations and gene-sets

We grouped variants into 35 sets (or masks) for gene-based tests based on variant annotations and allele frequencies. Variants were grouped based on 7 annotation categories: predicted loss of function (pLoF) variants, deleterious missense variants, possibly deleterious missense variants, all missense variants, pLoFs combined with each missense category. Variants were annotated using VEP^11^ using the variant from canonical transcripts. Variants annotated with stop gained, start lost, splice donor, splice acceptor, stop lost or frameshift were considered pLoFs. For missense variants, results from five prediction algorithms were used to determine their severity: SIFT^25^, PolyPhen2 HDIV, PolyPhen2 HVAR^26^, LRT^27^, and MutationTaster^28^. Variants were grouped into a deleterious missense category if predicted deleterious by all 5 algorithms, a possibly deleterious category if predicted deleterious by at least one algorithm, and an all missense category if not predicted deleterious by any algorithm. We considered five allele frequency bins: AAF < 1%, AAF < 0.5%, AAF < 0.1%, AAF < 0.001% and singletons.

For the BURDEN-ACAT test, we aggregated the p-values from the WST applied to the 35 masks. For the SKATO-ACAT and ACAT-V-ACAT tests we aggregated the p-values from SKAT-O and ACAT-V tests applied to 7 non-synonymous masks using a single AAF cutoff of 1% along with MAF-dependent variant weights. In each case aggregation was applied using ACAT (Cauchy combination method). The BURDEN-ACAT, SKATO-ACAT and ACAT-V-ACAT tests were then aggregated into a single GENE_P^19^ p-value using ACAT.

### Analysis and meta-analysis of UK Biobank

We analyzed WES data from the final release of the OQFE pipeline along with imputed genotypes for 469,376 samples in UK Biobank. Details of the exome sequencing^29^, phenotyping and array genotyping^30^, and imputation against TOPMed and ancestry assignment^1^, have been previously described. Both BMI and LDL were transformed using the rank-based inverse-normal approach. Association testing in REGENIE was performed using the covariates age, age^2^, sex, age by sex, exome batch, the top 10 array PCs, and the top 20 exome PCs.w2

### Description of ESM1v annotation methods

In the meta-analysis of UK Biobank, we tested 2 additional masks using ESM1v annotations to test for associations: ESM1v deleterious missense mutations predicted by the ESM1v model and ESM1v deleterious missense mutations combined with pLoFs. We downloaded the ESM-1v model from https://github.com/facebookresearch/esm. We computed the wildtype marginal score averaged across all five ESM-1v models. For proteins with sequences longer than 1022 amino acids, we centered the variant in the 1022 amino sequence. In cases where the variant was near the end of the protein, we included the maximal amount of variant amino acid context. Variants in the top 22% of missense scores were grouped into a “deleterious missense” category. We chose a 22% cutoff so that the same number of variants would be included in the deleterious missense category defined by the 5 prediction algorithms above as the ESM1v predictions.

## Data Availability

The study uses UK Biobank data that canbe accessed by approved researchers here https://www.ukbiobank.ac.uk
The software and code described in the paper is available at https://github.com/rgcgithub/remeta

## URLs

REGENIE https://rgcgithub.github.io/regenie/

metaSTAAR https://github.com/xihaoli/MetaSTAAR

UK Biobank RAP https://www.ukbiobank.ac.uk/enable-your-research/research-analysis-platform

AllofUs Researcher Workbench https://www.researchallofus.org/data-tools/workbench/

## Code availability

The C++ source code for REMETA is available from https://rgcgithub.github.io/remeta/ under an MIT License.

