## Supplementary Material for "Computationally efficient meta-analysis of gene-based tests using summary statistics in large-scale genetic studies"

November 8, 2024

**4 Supplementary Figures** **11**

**5 Supplementary Tables** **32**

### 1 Details of gene-based tests

Here we provide additional mathematical details of the gene-based tests implemented in REMETA. For each test the set up is as follows. Suppose we have phenotypes $y_i$ for $i = 1...n$ samples. Let $x_i \in \mathbb{R}^m$ be a vector of covariates (including the intercept) per sample and $X \in \mathbb{R}^{n\times m}$ a matrix with rows $x_i^T$. Let $g_i \in \{0, 1, 2\}^p$ be the vector counting the number of alternate alleles at each variant in a test and $G \in \mathbb{R}^{n\times p}$ a matrix with rows $g_i^T$. To keep notation consistent across sections individuals will be indexed from $i = 1...n$ and variants from $j = 1...p$.

#### 1.1 Burden testing

Our goal for burden testing in REMETA is to approximate meta-analysis of the default burden test in REGENIE, which uses collapsing variant test [1]. The collapsing variant test assumes that multiple damaging variants in the gene have the same impact as having just one. All the genotypes in the mask get collapsed into a single composite genotype:

$$C_i = \max\{g_{i1}, ..., g_{ip}\}$$

The max operator makes it challenging to reconstruct the collapsing burden test from single variant summary statistics. Instead, burden testing in REMETA is performed using a variation of the weighted sum test (WST) [2]. The WST takes a weighted sum of the variants in a mask to construct a composite genotype

$$C_i = \sum_{j=1}^{p} w_j g_{ij}$$

The $w_j$ are pre-specified variant weights. Here, each damaging variant in the mask is assumed to increases the magnitude of the effect on the phenotype. That effect can be weighted by different characteristics of the variant like its allele frequency. If the weights $w_j = 1$, and an individual has at most one damaging variant in the mask, then the collapsing variant test is equivalent to the WST. We refer to the special case of the WST with $w_j = 1$ as the sum test.

As most individuals are unlikely to have multiple rare damaging mutations in a single gene, we expect the composite genotypes for the collapsing variant test and the sum test to be similar. Indeed, p-values in from the sum test approximate p-values from the collapsing burden test (Supplementary Figure 6).

For both models association testing is performed using linear regression for quantitative traits and logistic regression for binary traits. For quantitative traits the model is

$$\begin{aligned} \eta_i &= x_i^T \alpha + C_i \beta \\ \mu_i &= \eta_i \\ y_i &\sim \mathcal{N}(\mu_i, \sigma^2) \end{aligned}$$

where $\sigma^2$ is the residual variance. For binary traits the model is

$$\begin{aligned} \eta_i &= x_i^T \alpha + C_i \beta \\ \mu_i &= \text{logistic}(\eta_i) \\ y_i &\sim \text{Bernoulli}(\mu_i) \end{aligned}$$

The benefit of the WST compared to the collapsing variant test is that a score test can be constructed from the score statistics of the individual variants and their covariance [3]

$$\begin{aligned} S &= G^T(y - \hat{\mu}) \\ \text{Cov}(S) &= G^T \text{Cov}(y - \hat{\mu}) G \end{aligned}$$

where $\hat{\mu}$ is estimated under the null hypothesis using either linear regression or logistic regression. The score statistic of the WST is given by $w^T S$, so we have

$$\begin{aligned} Q_{WST} &= (w^T S)^2 \\ \frac{Q_{WST}}{w^T \text{Cov}(S) w} &\sim \chi^2(1) \end{aligned}$$

##### 1.1.1 Estimating effect sizes

To estimate an effect size for the sum test, we use the observation that in the sum test each variant is assumed to have the same effect size. Specifically, if $\beta_1, ..., \beta_p$ are the effect sizes for the $p$ variants in a mask, then the sum test assumes $\beta := \beta_1 = \beta_2 = \cdots = \beta_p$. Thus, if $\hat{\beta}_1, ..., \hat{\beta}_p$ are the marginal estimates, then $\mathbb{E}[\hat{\beta}_j] = \beta$ for each variant $j$ in the mask. In principle, we could seek a convex combination $w_1, .., w_p$ of the effect sizes to construct and estimator for $\beta$. Let $\hat{\beta} = (\hat{\beta}_1, ...\hat{\beta}_j)^T$ and $w = (w_1, ..., w_p)^T$. Then we want to find

$$\begin{aligned} &\min_w \mathrm{Cov}(w^T \hat{\beta}) \\ \text{s.t.} \quad & w_j \geq 0 \text{ for } j = 1...p \\ &\sum_j w_j = 1 \end{aligned}$$

That is, we want to find a minimum variance unbiased estimator for $\beta$. If there is no correlation among the $\hat{\beta}$, then solution is equivalent to an inverse-variance weighted meta-analysis. As there is no analytic solution for this minimization problem, searching for a solution would require repeatedly solving a non-negative least squares problem for each burden mask—which is computationally intensive. Instead we use the estimate produced by inverse-variance weighted meta-analysis. This provides an unbiased estimate of the sum test effect size. If there is little LD among variants in the mask, this estimate should be close to the minimum variance estimator.

#### 1.2 The sequence kernel association test (SKAT)

Here we present a derivation for a special case of the SKAT and SKAT-O tests where the effect sizes follow a normal distribution. SKAT uses a variance component model where the effect sizes $\beta_j$ are the random effects. Let $\beta \in \mathbb{R}^p$, then the model we consider is

$$\begin{aligned} \beta &\sim N(0, \tau W) \\ W &:= \mathrm{diag}(w_1, ..., w_p) \end{aligned}$$

The $w_j$ are fixed weight parameters and $\tau$ is the variance component. For quantitative traits the phenotype is related to the genotypes by

$$\begin{aligned} \mu_i &= x_i^T \alpha + g_i^T \beta \\ y_i | \beta &\sim N\left(\mu_i, \sigma^2\right) \end{aligned}$$

where $\alpha$ are the fixed effects. For binary traits the SKAT model is

$$\begin{aligned} \mathrm{logit}(\mu_i) &= x_i^T \alpha + g_i^T \beta \\ y_i | \beta &\sim \mathrm{Bernoulli}(\mu_i) \end{aligned}$$

SKAT tests the null hypothesis $\tau = 0$ against the alternate hypothesis $\tau > 0$. We show in section 1.2.3 for quantitative traits and section 1.2.4 for binary traits that the test statistic for a score test is given by

$$Q_{SKAT} = (y - \hat{\mu})^T G W G^T (y - \hat{\mu})$$

where $\hat{\mu}$ is mean under the null hypothesis ($\tau = 0$).

##### 1.2.1 Computing the null distribution of $Q_{SKAT}$

We want to compute the null distribution of $Q_{SKAT}$. The terms $S := G^T(y - \hat{\mu})$ are the score statistics of the individual variants conditional on the estimate of $\alpha$. To compute their variance, note that in the limit of a large sample

$$\begin{bmatrix} G^T(y-\mu) \\ X^T(y-\mu) \end{bmatrix} \sim \mathcal{N}\left( \begin{bmatrix} 0 \\ 0 \end{bmatrix}, \begin{bmatrix} G^T V G & G^T V X \\ X^T V G & X^T V X \end{bmatrix} \right)$$

where $V = \sigma^2 I$ for quantitative traits and $V = \text{diag}\{\hat{\mu}_1(1-\hat{\mu_1}), ..., \hat{\mu}_n(1-\hat{\mu_n})\}$ for binary traits. At the maximum likelihood estimate of $\alpha$ we have $X^T(y-\mu) = 0$, so

$$\begin{aligned} G^T(y-\mu)\big|X^T(y-\mu) &\sim \mathcal{N}(0, G^TVG - G^TVX(X^TVX)^{-1}X^TVG) \\ &= \mathcal{N}(0, G^TPG) \\ P :&= V - VX(X^TVX)^{-1}X^TV \end{aligned}$$

Thus, under the null hypothesis ($\tau = 0$) we have

$$\begin{aligned} G^T(y-\hat{\mu}) &\sim \mathcal{N}\left(0, G^TP_0G\right) \\ P_0 &:= V - VX(X^TVX)^{-1}XV \end{aligned}$$

where $P_0$ is estimated under the null.

Now consider $\text{Var}(W^{1/2}S) = \text{Var}(W^{1/2}G^T(y-\hat{\mu})) = W^{1/2}G^TP_0GW^{1/2}$. The matrix is positive semi-definite, so it has an eigendecomposition $\Sigma\Lambda\Sigma^T$. Let

$$z = \Lambda^{-1/2}\Sigma^T\left(W^{1/2}G^T(y-\hat{\mu})\right)$$

and note that $z$ is $\mathcal{N}(0, I)$. Then we can write $Q_{SKAT}$ in terms of $z$

$$\begin{aligned} Q_{SKAT} &= (y-\hat{\mu})^TGWG^T(y-\hat{\mu}) \\ &= z^T\Lambda^{1/2}\Sigma^T\Sigma\Lambda^{1/2}z \\ &= z\Lambda z \\ &= \sum_{j=1}^{p}\lambda_j z_j^2 \end{aligned}$$

Each $z_j^2$ is a $\chi^2(1)$ random variable. The $\lambda_j$ are the eigenvalues of $W^{1/2}G^TP_0GW^{1/2}$. Thus $Q_{SKAT}$ has a mixture of chi-square distribution.

##### 1.2.2 Extension to SKAT-O

The SKAT model assumes the random effects $\beta_j$ are uncorrelated. If we instead test a model where the $\beta_j$ are correlated we get the model used by SKAT-O:

$$\begin{aligned} \beta &\sim N\left(0, \tau W^{1/2}\left((1-\rho)I + \rho\mathbf{1}\mathbf{1}^T\right)W^{1/2}\right) \\ W &= \text{diag}(w_1, .., w_p) \end{aligned}$$

The term $\rho$ is the correlation between each pair of $\beta_k$ and $\beta_{k'}$. Let $R_\rho = (1-\rho)I + \rho\mathbf{1}\mathbf{1}^T$. Replacing $\tau W$ by $\tau W^{1/2}R_\rho W^{1/2}$ in the derivations in sections 1.2.3 and 1.2.4 gives us the score statistic for the variance component test

$$Q_\rho = (y-\hat{\mu})^TGW^{1/2}R_\rho W^{1/2}G^T(y-\hat{\mu})$$

We can expand $Q_\rho$ into a weighted sum of a burden test and $Q_{SKAT}$

$$\begin{aligned} Q_\rho &= (y-\hat{\mu})^TGW^{1/2}((1-\rho)I + \rho\mathbf{1}\mathbf{1}^T)W^{1/2}G^T(y-\hat{\mu}) \\ &= (1-\rho)(y-\hat{\mu})^TGW^{1/2}W^{1/2}G^T(y-\hat{\mu}) + \rho(y-\hat{\mu})GW^{1/2}\mathbf{1}\mathbf{1}^TW^{1/2}G^T(y-\hat{\mu}) \\ &= (1-\rho)\sum_{j=1}^{p}\left(w_j^{1/2}\sum_{i=1}^{n}g_{ij}(y_i-\hat{\mu}_i)\right)^2 + \rho\left(\sum_{j=1}^{p}w_j^{1/2}\sum_{i=1}^{n}g_{ij}(y_i-\hat{\mu}_i)\right)^2 \\ &= (1-\rho)\left(\sum_{j=1}^{p}w_j^{1/2}S_j^2\right) + \rho\left(\sum_{j=1}^{p}w_j^{1/2}S_j\right)^2 \\ &= (1-\rho)Q_{SKAT} + \rho Q_{BURDEN} \end{aligned}$$

##### 1.2.3 Computing the score statistic for a quantitative trait

The score statistic is defined to be the derivative of the log likelihood. Thus we first need to compute

$$L(\alpha, \tau) := p(y) = \int p(y|\beta)p(\beta)\, d\beta = \int N(y|X\alpha + G\beta, \sigma^2 I)N(\beta|0, \tau W)\, d\beta$$

The computation for marginalizing a product of Gaussians is well known (e.g. see [4]) The result is

$$p(y) = N\left(y|X\beta, \sigma^2 I + \tau GWG^T\right)$$

The log-likelihood of $y$ is

$$l(\alpha, \tau) = -\frac{1}{2}(y - X\alpha)^T \left(\sigma^2 I + \tau GWG^T\right)^{-1}(y - X\alpha) - \frac{1}{2}\log\left|\sigma^2 I + \tau GWG^T\right| + \text{const wrt } \tau$$

Taking derivatives with respect to $\tau$ gives us the score statistic. For the first term write out the quadratic form as a sum, use $\frac{\partial}{\partial x}A^{-1} = -A^{-1}(\frac{\partial}{\partial x}A)A^{-1}$, then rewrite as a quadratic form. For the second term use $\frac{\partial}{\partial x}\log|A| = \text{Tr}(A^{-1}(\frac{\partial A}{\partial x}))$.

$$\begin{aligned}\frac{\partial l}{\partial \tau} &= \frac{1}{2}(y - X\alpha)^T (\sigma^2 I + \tau GWG^T)^{-1} GWG^T (\sigma^2 I + \tau GWG^T)^{-1}(y - X\alpha) \\ &\quad - \frac{1}{2}\text{Tr}\left(\left(\sigma^2 I + \tau GWG^T\right)^{-1} GWG^T\right)\end{aligned}$$

Under the null hypothesis $H_0 : \tau = 0$ we have

$$\frac{1}{2\sigma^4}(y - X\alpha)^T GWG^T (y - X\alpha) - \frac{1}{2}\text{Tr}\left((\sigma^2 I)^{-1} + GWG^T\right)$$

Note that only the first term is random while the second term is constant. Therefore to test the null hypothesis we only need the first term. Rescaling by $2\sigma^4$ gives us the SKAT test statistic

$$Q = (y - \mu)^T GWG^T (y - \mu)$$

##### 1.2.4 Computing the score statistic for a binary trait

Similar to above, we want to compute

$$L(\alpha, \tau) := p(y) = \int p(y|\beta)p(\beta)\, d\beta = \int \prod_{i=1}^{n} \text{Bernoulli}(y_i|\mu_i)N(\beta|0, \tau W)\, d\beta$$

Unlike above we cannot compute the integral in closed form. Instead we use an argument similar to [5]. To make this problem tractable we need to take two Taylor approximations. First approximate $p(y|\beta) = \exp\{\log p(y|\beta)\}$ using a Taylor expansion around $\beta = 0$, the hypothesis we want to test. Taking gradients with respect to $\beta$

$$\begin{aligned}\log p(y|\beta) &= \sum_{i=1}^{n} y_i \log \mu_i + (1 - y_i)\log(1 - \mu_i) \\ \implies \nabla_\beta \log p(y|\beta) &= \sum_{i=1}^{n} y_i \frac{1}{\mu_i}\mu_i(1 - \mu_i)g_i - (1 - y_i)\frac{1}{1 - \mu_i}\mu_i(1 - \mu_i)g_i \\ &= \sum_{i=1}^{n}(y_i - \mu_i)g_i \\ &= G^T(y - \mu) \\ \implies \nabla\nabla_\beta \log p(y|\beta) &= G^T V G \\ V &:= \text{diag}(\mu_1(1 - \mu_1), ..., \mu_n(1 - \mu_n))\end{aligned}$$

Then we can compute

$$\nabla_\beta \exp\{\log p(y|\beta)\} = \exp\{\log p(y|\beta)\}\nabla_\beta \log p(y|\beta)$$
$$\nabla\nabla_\beta \exp\{\log p(y|\beta)\} = \exp\{\log p(y|\beta)\}\left(\nabla_\beta \log p(y|\beta)\nabla_\beta \log p(y|\beta)^T + \nabla\nabla_\beta \log p(y|\beta)\right)$$

Expanding around $\beta = 0$

$$\exp\{\log p(y|\beta)\} \approx \exp\{\log p(y|\beta = 0)\}\left(1 + (y-\mu)^T G\beta + \frac{1}{2}\beta^T\left(G^T(y-\mu)(y-\mu)^T G + G^T V G\right)\beta\right)$$

where $\text{logit}(\mu) = X\beta$. Then we can write the integral as an expectation

$$L(\alpha,\tau) = E_\beta\left[\exp\{\log p(y|\beta)\}\right] \approx p(y|\beta = 0)\left(1 + \frac{1}{2}\text{Tr}(\tau G^T(y-\mu)(y-\mu)^T GW + \tau G^T V G W)\right)$$

Taking another Taylor expansion around $\tau = 0$ of the log likelihood

$$\begin{aligned} l(\alpha,\tau) := \log p(y) &= \log p(y|\beta = 0) + \log\left(1 + \frac{\tau}{2}\text{Tr}(G^T(y-\mu)(y-\mu)^T GW + G^T V G W)\right) \\ &\approx \log p(y|\beta = 0) + \frac{\tau}{2}\text{Tr}\left(G^T(y-\mu)(y-\mu)^T GW + G^T V G W\right) \end{aligned}$$

Rearranging terms using the "trace trick"

$$\begin{aligned} l(\alpha,\tau) &\approx \log p(y|\beta = 0) + \frac{\tau}{2}\text{Tr}(G^T(y-\mu)(y-\mu)^T GW) + \frac{\tau}{2}\text{Tr}(G^T V G W) \\ &= \log p(y|\beta = 0) + \frac{\tau}{2}(y-\mu)^T GWG^T(y-\mu) + \frac{\tau}{2}\text{Tr}(G^T V G W) \end{aligned}$$

Now we can compute the score statistic as before

$$\frac{\partial l(\alpha,\tau)}{\partial\tau} = \frac{\partial}{\partial\tau}\log p(y) = \frac{1}{2}(y-\mu)^T GWG^T(y-\mu) + \frac{1}{2}\text{Tr}(G^T V G W)$$

Again as before, the first term is the only random term. Rescaling by $\frac{1}{2}$ gives us the result

$$Q = (y-\mu)^T GWG^T(y-\mu)$$

#### 1.3 Aggregated Cauchy association test (ACAT-V)

The aggregated Cauchy association test (ACAT) is a method for combining p-values across multiple tests [6]. Similar to Fisher's method which transforms p-values to $\chi^2$ random variables, or Stouffer's method which transforms p-values to $Z$ scores, ACAT transforms $p$-values to Cauchy distributed random variables. It uses the fact that under some mild assumptions, convex combinations of Cauchy transformed $p$-values will follow a standard Cauchy distribution — even if the $p$-values are correlated. If $p_1, ..., p_d$ are the $p$-values to be combine and $w_1, ..., w_d$ the weights, then the test statistic is

$$T = \sum_{i=1}^{d} w_i \tan\{(0.5 - p_i)\pi\} \sim \text{Cauchy}(0,1)$$

When the weights are all equal ACAT can be used to combine p-values from correlated tests to correct for multiple testing. In the context of rare variant association testing, p-values can be combined into a set-based test by grouping variables in an annotation category [7]. Like the WST and SKATO, per variant weights can be chosen based on features of the variant like its minor allele frequency. If $w_j$ is the weight for a variant $j$ in WST or SKATO, then for ACAT-V others have suggested [7] to set weight to be

$$\begin{aligned} w_{j,ACAT-V} &= w_j^2 f_j(1-f_j)\bar{w} \\ \bar{w} &= \sum_j w_{j,ACAT-V} \end{aligned}$$

Thus

$$T_{ACAT-V} = \frac{1}{\bar{w}}\sum_{j=1}^{p} w_{j,ACAT-V}\tan\{(0.5 - p_j)\pi\}$$

#### 2 Saddlepoint approximation for unbalanced binary traits

For unbalanced binary using a normal approximation to compute the distribution of the test statistic can lead to inflated type I error. In this setting, saddlepoint approximation (SPA) [8] has been shown to help control type I error [9]. SPA uses the cumulant-generating function of the test statistic to approximate its distribution. The test statistic for a score test in logistic regression is

$$S_j = \sum_{i=1}^{n} g_{ij}(y_i - \hat{\mu_i})$$

The cumulant generating function is

$$\begin{aligned} K(t) &= \log\left(\mathbb{E}_{H_0}[e^{tS_j}]\right) \\ &= \sum_{i=1}^{n} \log(1 - \hat{\mu_i} + \hat{\mu_i} e^{g_{ij}t}) - t \sum_{i=1}^{n} g_{ij}\hat{\mu_i} \end{aligned}$$

where $\hat{\mu_i}$ is estimated under the null hypothesis. The first two derivatives of $K(t)$ are

$$\begin{aligned} K'(t) &= \sum_{i=1}^{n} \frac{\hat{\mu_i} g_{ij}}{1 - \hat{\mu_i} + \hat{\mu_i} e^{g_{ij}t}} - \sum_{i=1}^{n} g_{ij}\hat{\mu_i} \\ K''(t) &= \frac{\hat{\mu_i}(1 - \hat{\mu_i}) g_{ij}^2 e^{-g_{ij}t}}{\left((1 - \hat{\mu_i}) e^{-g_{ij}t} + \hat{\mu_i}\right)^2} \end{aligned}$$

Then the saddlepoint approximation to the distribution of $S_j$ is given by

$$\begin{aligned} \Pr(S_j < s) &= \Phi\left(w + \frac{1}{w}\log\left(\frac{v}{w}\right)\right) \\ w &= \text{sign}(\delta^*)\sqrt{2(\delta^* s - K(\delta^*)} \\ v &= \delta^* \sqrt{K''(\delta^*)} \end{aligned}$$

where $\Phi$ is the standard normal distribution and $\delta^*$ is the solution to $K'(t) = s$.

When only an intercept is included in the model, a SPA can be computed from case-control counts and genotype counts alone. The intercept is given by $\hat{\mu_i} = \hat{\mu} = \frac{\text{\# cases}}{\text{\# cases+\# controls}}$. If we let $n_{ref}$, $n_{het}$ and $n_{alt}$ be the number of individuals with homozygous reference, heterzygous, and homozygous alternate genotypes respectively, we have

$$K(t) = \sum_{x \in \{ref, het, alt\}} n_x \log(1 - \hat{\mu} + \hat{\mu} e^{g_x t}) \tag{1}$$

where $g_x$ are the (possibly centered and standardized) reference, heterozygous, and homozygous alternate genotypes. This approach has been shown to work well in the setting of single-variant meta-analysis [10].

To extend to gene-based tests, we use the observation the sum test approximates the collapsing burden test, and that we can compute genotype counts from the collapsing burden test. Let $n^s_{cases}$ and $n^s_{controls}$ be the number of cases and controls for a study $s$. Furthermore, let $g^s_x$ be the number of genotype counts for $x \in \{ref, het, alt\}$. We compute

$$\begin{aligned} \hat{\mu} &= \frac{\sum_s n^s_{cases}}{\sum_s n^s_{cases} + n^s_{controls}} \\ n_x &= \sum_s n^2_x \text{ for } x \in \{ref, het, alt\} \end{aligned}$$

We then use cumulant generating function in equation 1 to compute a p-value for the sum test.

For the WST and SKAT-O, we use the burden $p$-value computed by SPA to compute a calibration factor similar to [11]. Specifically, let $p_{ST}$ and $p_{SPA}$ be the p-values computed from the sum test and SPA respectively. Then

$$r = \max\left(1, \frac{\chi^2_{quantile}(1 - p_{ST})}{\chi^2_{quantile}(1 - p_{SPA})}\right)$$

We use $r$ as a calibration factor to increase the covariance of the score statistics by replacing $\text{Cov}(S)$ with $r\text{Cov}(S)$.

### 3 Estimating mask genotype counts and allele frequencies

#### 3.1 Preliminaries

We start with a result that will be useful for estimating genotype counts of burden masks. Let $G_1 = X_1^{(1)} + X_1^{(2)}$ and $G_2 = X_2^{(1)} + X_2^{(2)}$ be two genotypes made up of the variants $X_j^{(h)}$ on haplotypes $h \in \{1, 2\}$. We want to compute $\Pr(X_1^{(h)} = 1, X_2^{(h)} = 1)$ from the LD between $G_1$ and $G_2$.

Assuming each haplotype is inherited independently, then $\mathbb{E}\left[X_1^{(h)} X_2^{(k)}\right] = \mathbb{E}\left[X_1^{(h)}\right]\mathbb{E}\left[X_1^{(k)}\right]$ for $h \neq k$. Let $f_1$ and $f_2$ be the frequencies of variants 1 and 2. We have

$$\begin{aligned}
\text{Cov}(G_1, G_2) &= \text{Cov}\left(X_1^{(1)} + X_1^{(2)}, X_2^{(1)} + X_2^{(2)}\right) \\
&= \mathbb{E}\left[\left(X_1^{(1)} + X_1^{(2)}\right)\left(X_2^{(1)} + X_2^{(2)}\right)\right] - \mathbb{E}\left[X_1^{(1)} + X_1^{(2)}\right]\mathbb{E}\left[X_2^{(1)} + X_2^{(2)}\right] \\
&= \mathbb{E}\left[X_1^{(1)} X_2^{(1)}\right] + \mathbb{E}\left[X_1^{(1)} X_2^{(2)}\right] + \mathbb{E}\left[X_1^{(2)} X_2^{(1)}\right] + \mathbb{E}\left[X_1^{(2)} X_2^{(2)}\right] - 4f_1 f_2 \\
&= \mathbb{E}\left[X_1^{(1)} X_2^{(1)}\right] + \mathbb{E}\left[X_1^{(1)}\right]\mathbb{E}\left[X_2^{(2)}\right] + \mathbb{E}\left[X_1^{(2)}\right]\mathbb{E}\left[X_2^{(1)}\right] + \mathbb{E}\left[X_1^{(2)} X_2^{(2)}\right] - 4f_1 f_2 \\
&= \mathbb{E}\left[X_1^{(1)} X_2^{(1)}\right] + \mathbb{E}\left[X_1^{(2)}\right]\mathbb{E}\left[X_2^{(1)}\right] - 2f_1 f_2 \\
&= 2\mathbb{E}\left[X_1^{(1)} X_2^{(1)}\right] - 2f_1 f_2 \\
&= 2\mathbb{E}\left[X_1 X_2\right] - 2f_1 f_2 \\
\implies \Pr(X_1 = 1, X_2 = 1) = \mathbb{E}[X_1 X_2] &= \frac{1}{2}\text{Cov}(G_1, G_2) + f_1 f_2
\end{aligned}$$

#### 3.2 Genotype counts

We derived an estimator for the genotype counts of a burden mask from the genotype counts of the variants in the mask and their LD. Let $G_{ji}$ be the genotype of variant $j$ in individual $i$. Let $N_{G_j=1}$ and $N_{G_j=2}$ be the heterozygous and homozygous genotype counts respectively. We want to find some combination of the $N_{G_j=1}$ and $N_{G_j=2}$ to estimate the heterozygote and homozygote genotype counts of the mask. Focusing on heterozygotes (the homozygous case is similar), if $Y_i = \max\{G_{1i}, ..., G_{pi}\}$ is the mask genotype we want to find coefficients $c_m$ that

$$N_{Y_i=1} = \sum_{j=1}^{p} c_j N_{G_j=1}$$

One possible strategy is to compute the $c_j$ sequentially. If we set $c_1 = 1$, then for each $c_{j+1}$ we can compute

$$\begin{aligned}
\Pr(G_1 \neq 1, G_2 \neq 1, ..., G_j \neq 1, G_{j+1} = 1) &= \Pr(G_1 \neq 1, G_2 \neq 1, ..., G_j \neq 1 | G_{j+1} = 1)\Pr(G_{j+1} = 1) \\
&= \Pr(G_1 \neq 1, G_2 \neq 1, ..., G_j \neq 1 | G_{j+1} = 1)\frac{N_{G_j=1}}{N}
\end{aligned}$$

This gives us the proportion of heterozygotes at $G_{j+1}$ that we have not already counted among $G_1, ..., G_j$. The coefficient we want it

$$c_{j+1} = \Pr(G_1 \neq 1, G_2 \neq 1, ..., G_j \neq 1 | G_{j+1} = 1)$$

If we assume that all LD among $G_1, ..., G_j$ is explained by $G_{j+1}$, then we can approximate

$$\Pr(G_1 \neq 1, G_2 \neq 1, ..., G_j \neq 1 | G_{j+1} = 1) \approx \prod_{m=1}^{j} \Pr(G_m \neq 1 | G_{j+1} = 1)$$

Let $f_j$ be the allele frequency of variant $j$, and suppose that $X_j^{(1)} = 1$ and $X_j^{(2)} = 0$ are the haplotypes for variant $j$ (the labeling of the haplotype is arbitrary). We have

$$\Pr(G_m = 1 | G_j = 1) = \Pr(X_m^{(1)} = 1 | X_j^{(1)} = 1)(1 - f_j) + (1 - \Pr(X_m^{(1)} = 1 | X_j^{(1)} = 1)) f_j$$

The first term is the case where $G_j = 1$ and $G_m = 1$ along the same haplotype, the second term is when $G_j = 1$ and $G_m = 1$ along different haplotypes. This gives us

$$\Pr(G_m \neq 1 | G_j = 1) = 1 - \Pr(G_m = 1 | G_j = 1)$$

The argument for homozygotes is similar. We want to compute

$$\begin{aligned} \Pr(G_m = 2 | G_j = 2) &= \Pr(X_m^{(1)} = 1 | X_j^{(1)} = 1) \Pr(X_m^{(2)} = 1 | X_j^{(2)} = 1) \\ &= \Pr(X_m^{(1)} = 1 | X_j^{(1)} = 1)^2 \end{aligned}$$

So

$$\Pr(G_j \neq 2 | G_m = 2) = 1 - \Pr(G_j = 2 | G_m = 2)$$

#### 3.3 Estimating allele frequencies when genotype counts are unavailable

When genotype counts are unavailable, we can estimate the allele frequency of burden masks from the allele frequencies of variants in the mask and their LD. To derive the estimator we model the haplotype of the variants contributing to the mask. Suppose $X_1, ..., X_p$ where $X_j \in \{0, 1\}$ are the variants in the haplotype constributing to the mask. The mask haplotype is $Y = \max\{X_1, ..., X_p\}$. The frequency of the haplotype is

$$\begin{aligned} \mathbb{E}[Y] &= \Pr(Y = 1) = 1 - \Pr(Y = 0) \\ &= 1 - \Pr(X_1 = 0, ..., X_p = 0) \end{aligned}$$

If the mask is in Hardy-Weinberg equilibrium, then the frequency of the mask haplotype is equivalent to the frequency of the mask. We want to approximate $\Pr(X_1 = 0, ..., X_p = 0)$ from something computable from the LD matrix. If we model the variants in the mask as a Markov chain, then this term becomes

$$\Pr(X_1 = 0, ..., X_p = 0) \approx \Pr(X_1 = 0) \prod_{t=2}^{T} \Pr(X_t = 0 | X_{t-1} = 0)$$

The motivation for this choice is that $\Pr(X_t = 0 | X_{t-1} = 0)$ can be computed from the LD matrix. Specifically

$$\begin{aligned} \Pr(X_t = 0 | X_{t-1} = 0) &= \frac{\Pr(X_t = 0, X_{t-1} = 0)}{f_{t-1}} \\ &= \frac{1}{f_{t-1}} \left(1 - \Pr(X_t = 1) - \Pr(X_{t-1} = 1) + \Pr(X_t = 1, X_{t-1} = 1)\right) \end{aligned}$$

What remains is to pick an order to evaluate the Markov chain. We start by setting $X_1$ to the most frequent variant. For each subsequent variant we choose the variant in the most LD with the previous. Thus for $X_2$, we choose the variant with the most LD to $X_1$.

### 4 Supplementary Figures

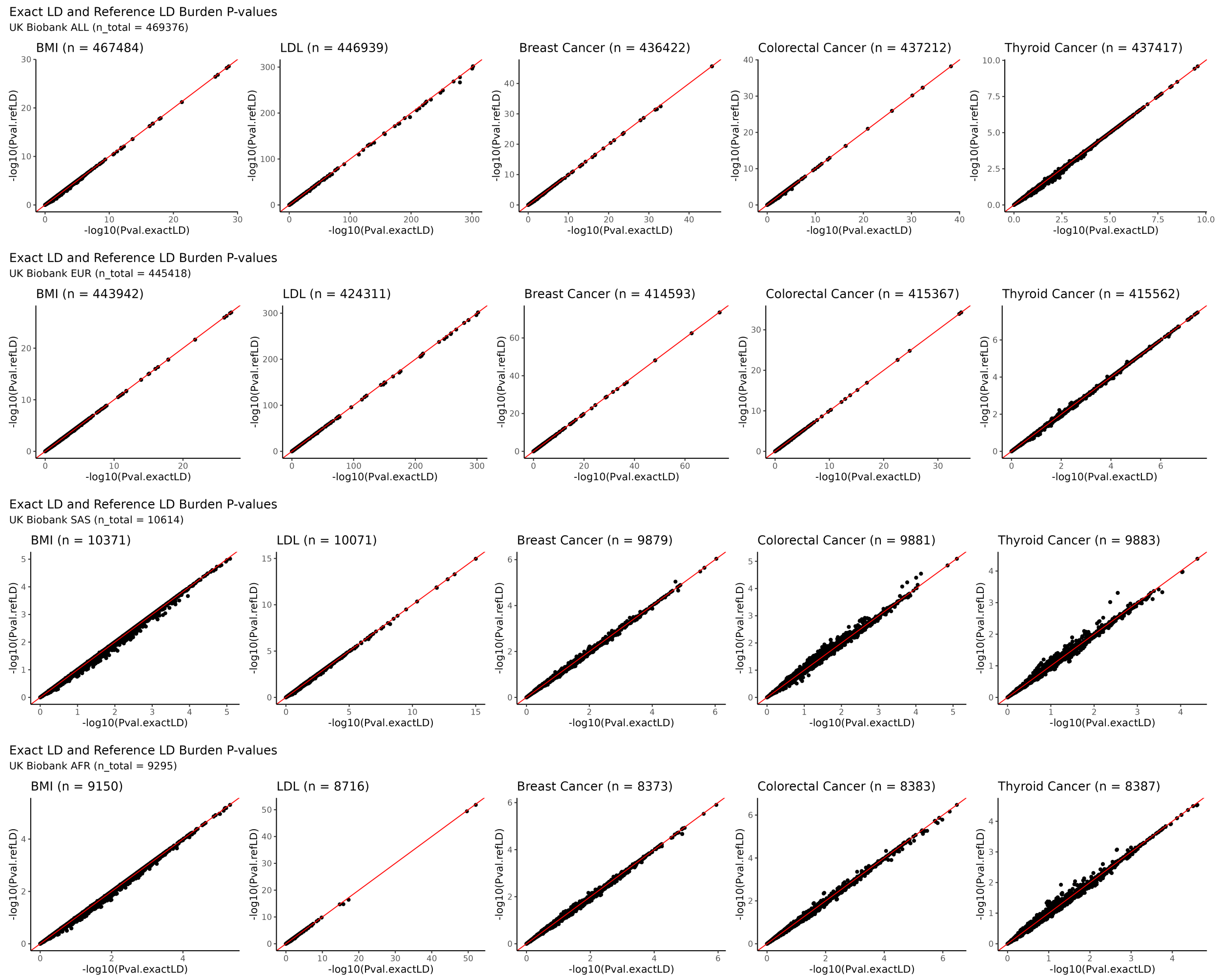

Supplementary Figure 1: **Scatterplots comparing $p$-values of burden tests computed using the exact covariance of the score statistics and the approximation using a reference LD panel.** Each row corresponds to a cohort, and each column corresponds to trait. The panels compare $p$-values computed using the covariance of the score statistics in the sample ($x$-axis; Pval.exactLD; sample sizes given above each panel) to $p$-values computed using a reference LD panel computed in the cohort ($y$-axis; Pval.refLD; sample sizes given by n_total). Burden tests are computed across 7 annotations categories at 5 allele frequency bins, resulting in 35 points each scatterplot for each gene.

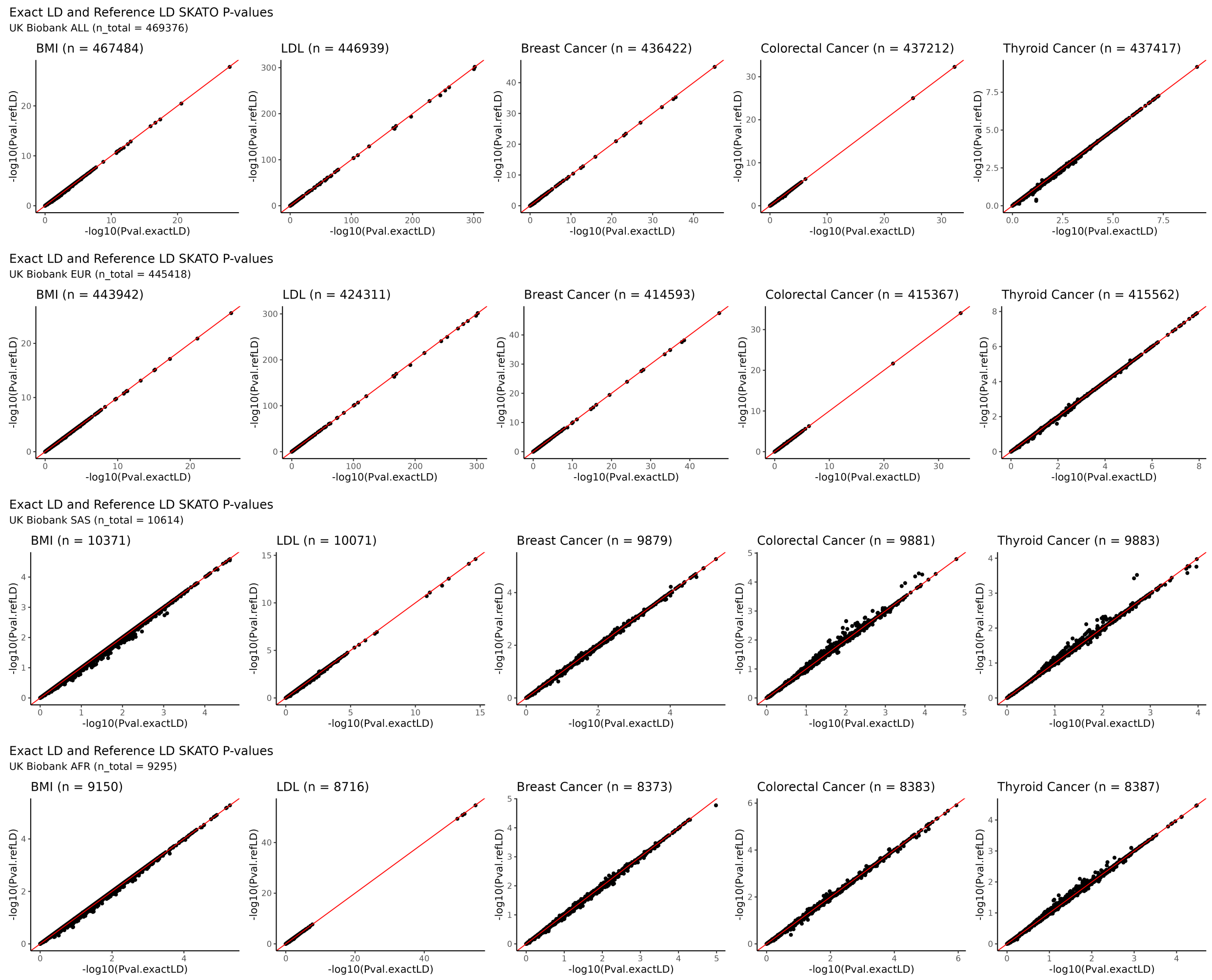

Supplementary Figure 2: **Scatterplots comparing SKAT-O $p$-values computed using the exact covariance of the score statistics and the approximation using a reference LD panel.** Each row corresponds to a cohort, and each column corresponds to trait. The panels compare $p$-values computed using the covariance of the score statistics in the sample ($x$-axis; Pval.exactLD; sample sizes given above each panel) to $p$-values computed using a reference LD panel computed in the cohort ($y$-axis; Pval.refLD; sample sizes given by n_total). SKAT-O is computed across 7 annotation categories using a 1% AAF cutoff, resulting in 7 points in each scatterplot for each gene.

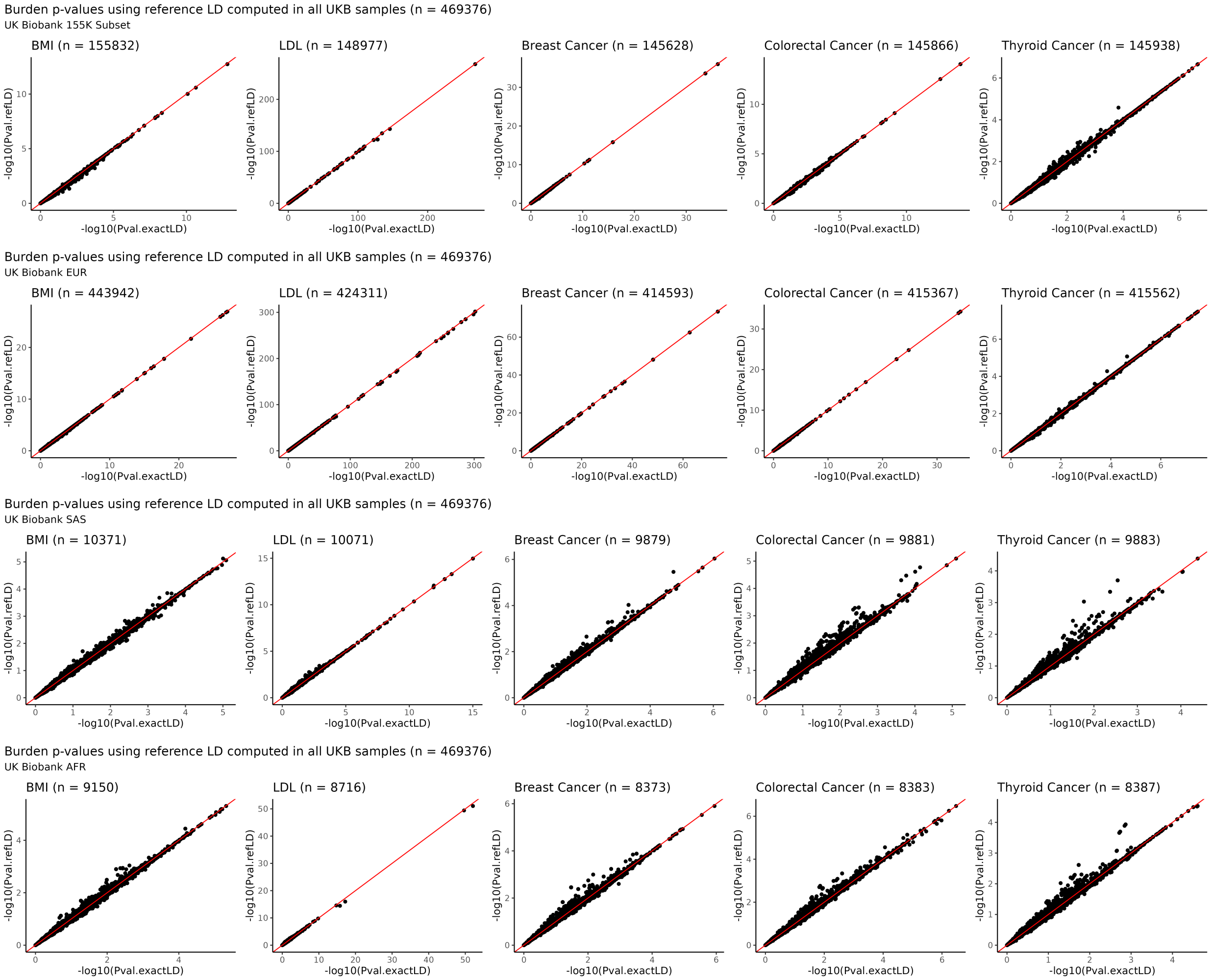

Supplementary Figure 3: **Scatterplots comparing $p$-values of burden tests computed using the exact covariance of the score statistics and a reference LD matrix computed in UK Biobank ALL.** The panels compare $p$-values computed using the covariance of the score statistics in the sample ($x$-axis; Pval.exactLD; sample sizes given above each panel) to $p$-values computed using a reference LD panel computed in all samples in UK Biobank ($y$-axis; Pval.refLD; sample sizes given by n_total).

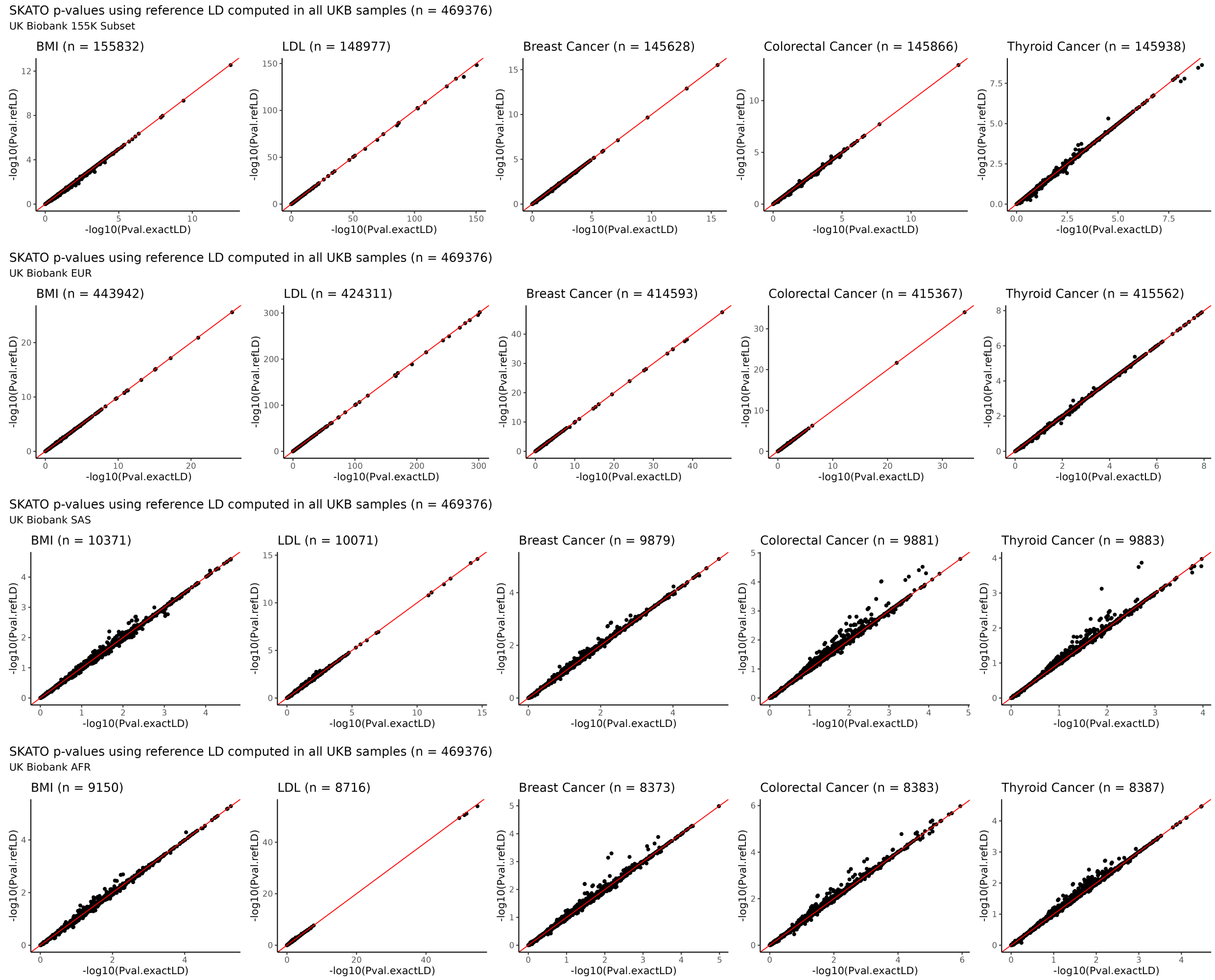

Supplementary Figure 4: **Scatterplots comparing SKAT-O $p$-values computed using the exact covariance of the score statistics and a reference LD matrix computed in UK Biobank ALL.** The panels compare $p$-values computed using the covariance of the score statistics in the sample ($x$-axis; Pval.exactLD; sample sizes given above each panel) to $p$-values computed using a reference LD panel computed in all samples in UK Biobank ($y$-axis; Pval.refLD; sample sizes given by n_total).

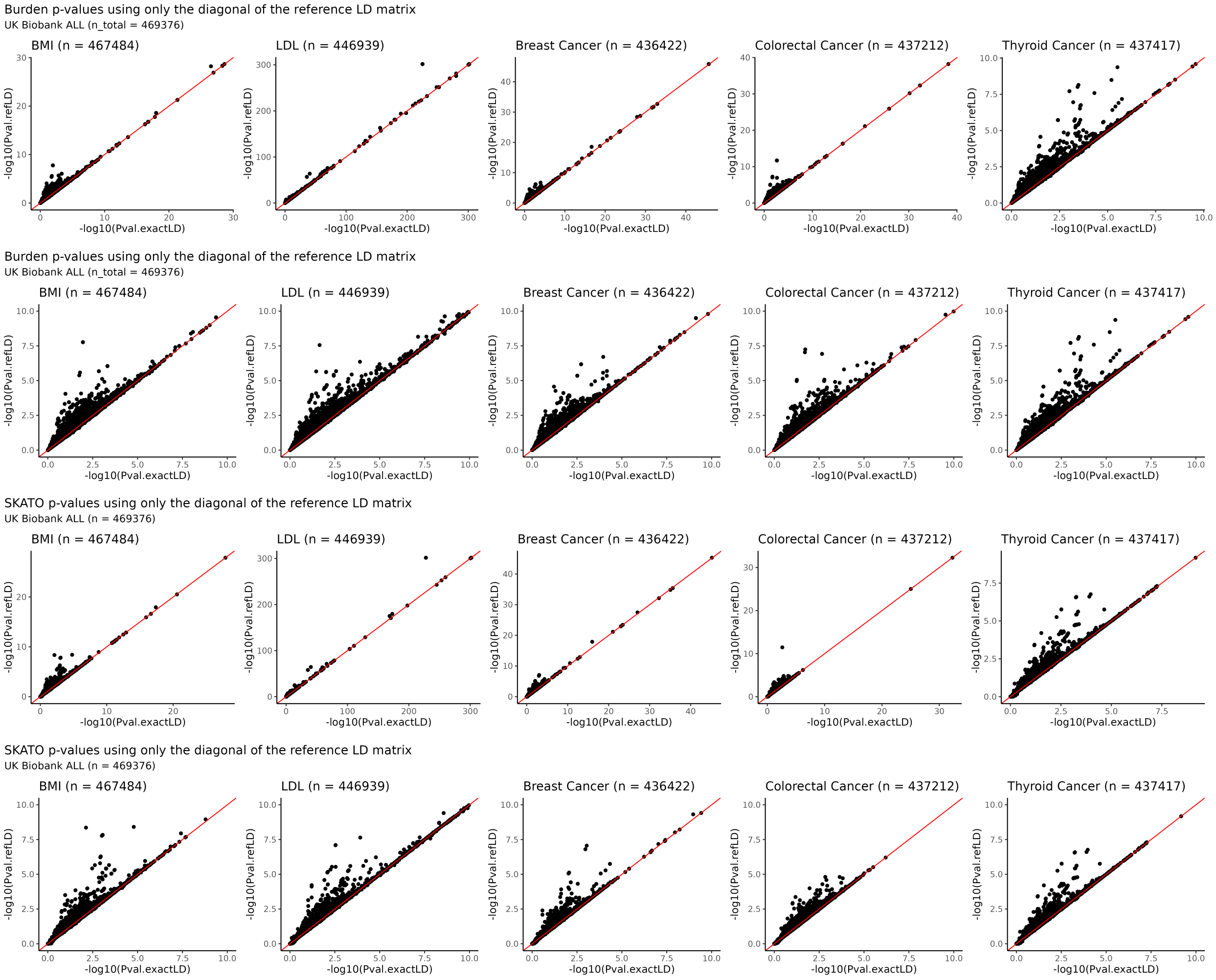

Supplementary Figure 5: **Scatterplots comparing burden test and SKAT-O $p$-values computed using the exact covariance of the score statistics $p$-values computed by ignoring LD between variants.** The top two rows display $p$-values for burden tests (note the change in scale between rows 1 and 2), and the bottom two rows display $p$-values for SKAT-O. The panels compare $p$-values computed using the covariance of the score statistics in the sample ($x$-axis; Pval.exactLD; sample sizes given above each panel) to $p$-values computed using only the diagonal of the covariance ($y$-axis; Pval.refLD).

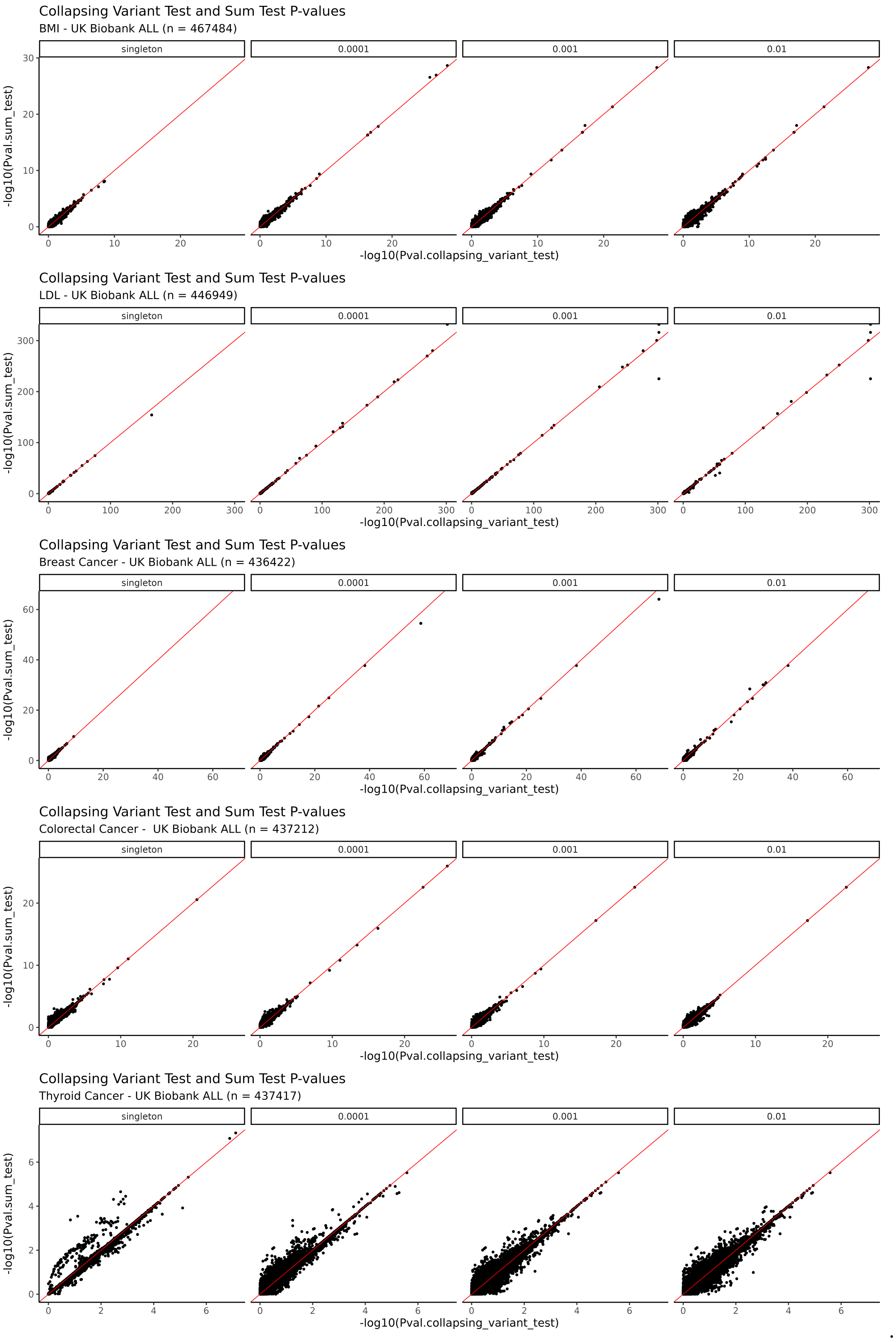

Supplementary Figure 6: **Scatterplot comparing $p$-values from the collapsing variant test and the sum test across 5 traits in UK Biobank ALL.** Each row corresponds to a trait, and each column corresponds to an AAF bin for a burden test. Each panel includes burden masks computed across 7 annotation categories. Burden testing for the collapsing variant test and sum test was performed in REGENIE using the –build-mask max and –build-mask sum options respectively.

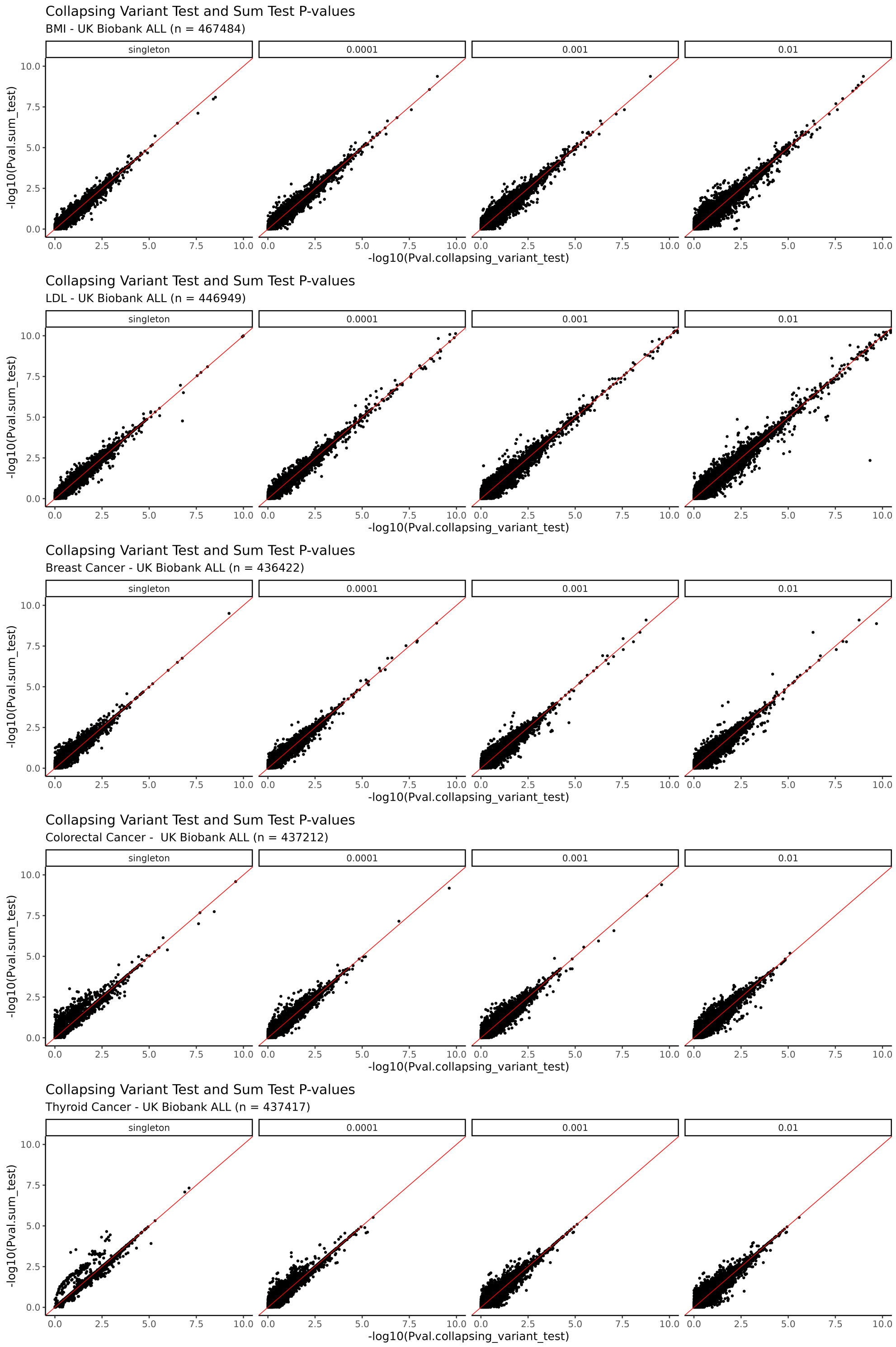

Supplementary Figure 7: **Zoomed in scatterplot comparing *p*-values from the collapsing variant test and the sum test.** Each row corresponds to a trait, and each column corresponds to an AAF bin for a burden test. Each panel includes burden masks computed across 7 annotation categories for each gene. Burden testing for the collapsing variant test and sum test was performed in REGENIE using the –build-mask max and –build-mask sum options respectively.

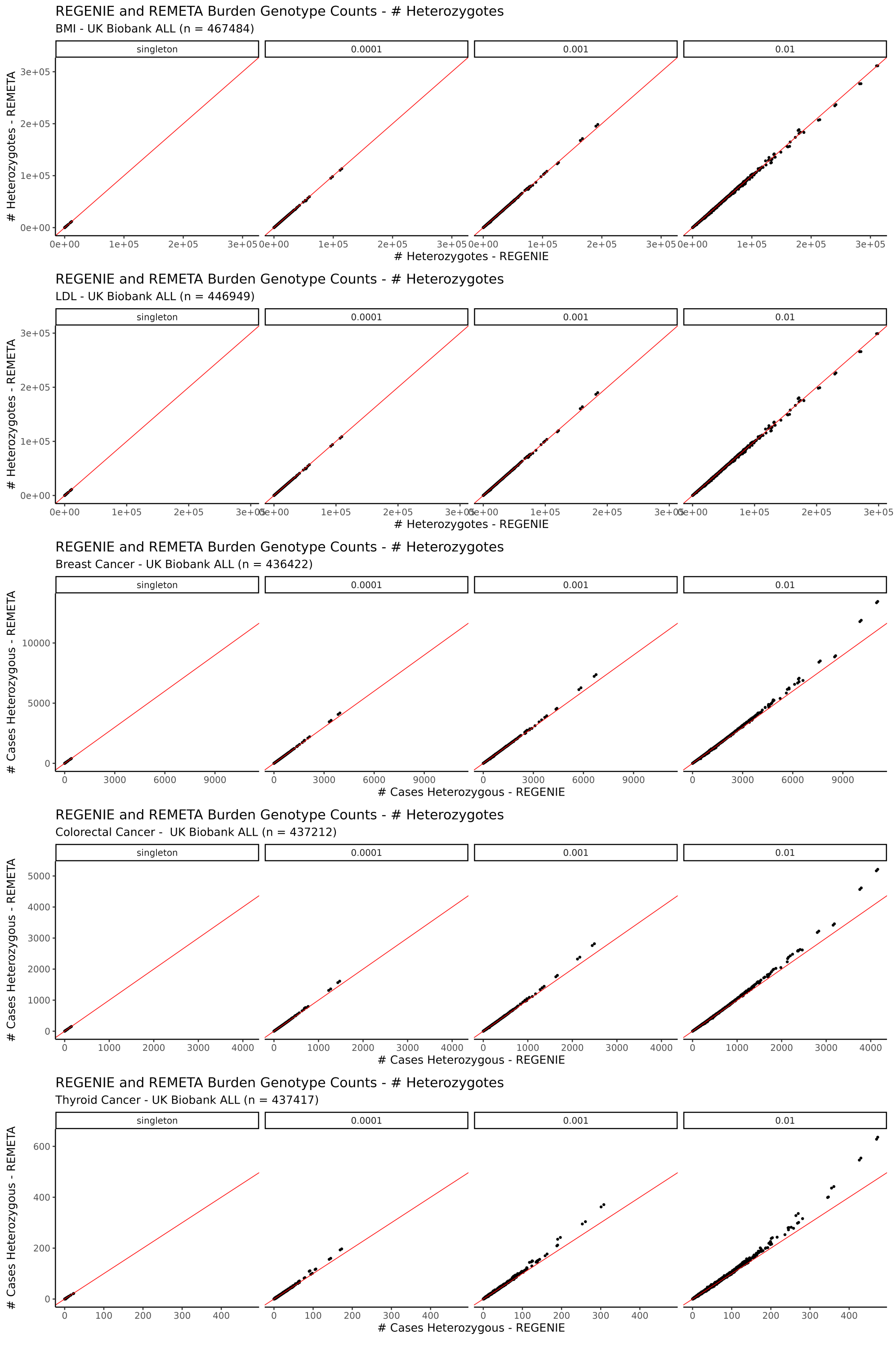

Supplementary Figure 8: **Scatterplot comparing genotype counts of burden masks computed by REGENIE and REMETA.** Each row corresponds to a trait, and each column corresponds to an AAF bin for a burden test. Each panel includes burden masks computed across 7 annotation categories. For quantitative traits, genotypes counts in the whole sample are displayed. For binary traits, genotype counts among cases are displayed.

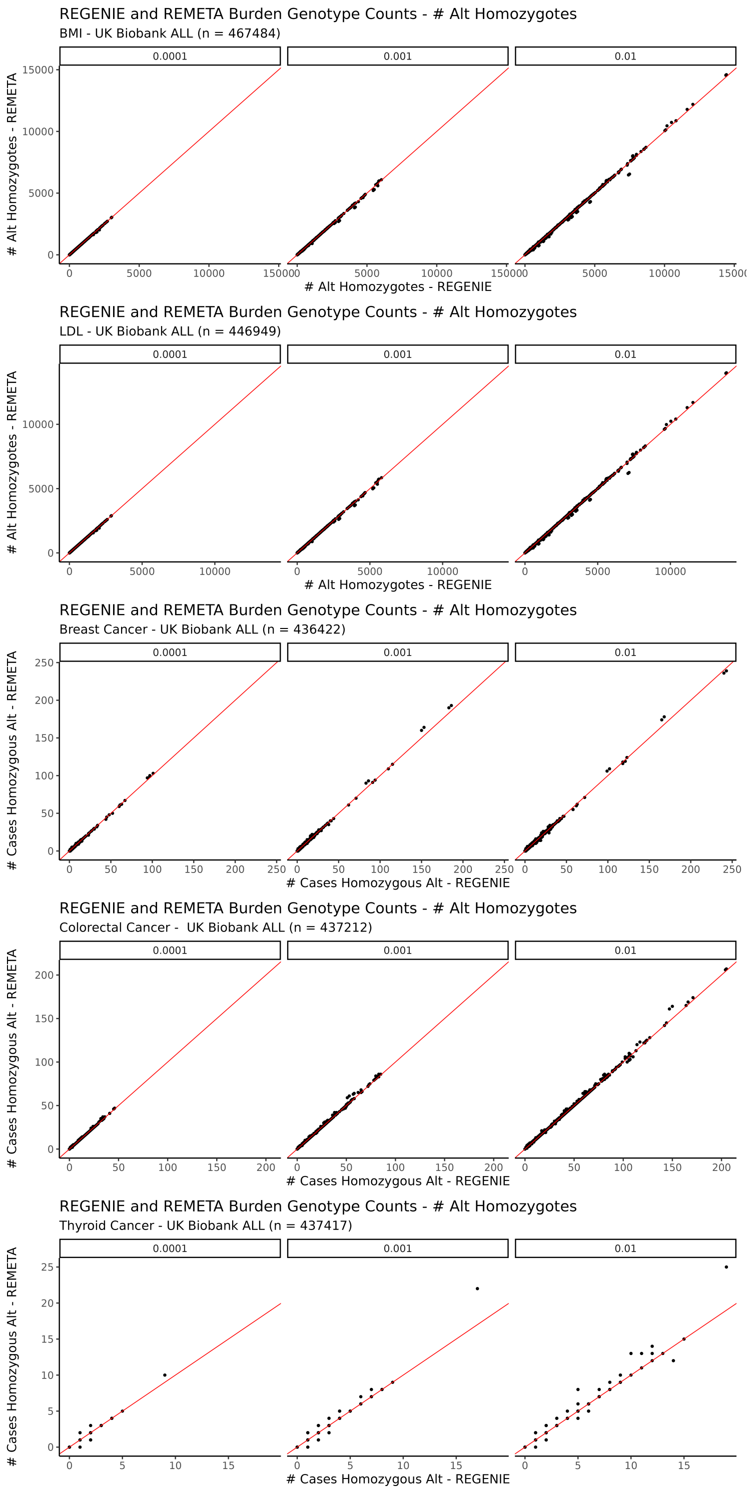

Supplementary Figure 9: **Scatterplot comparing genotype counts of burden masks computed by REGENIE and REMETA.** Each row corresponds to a trait, and each column corresponds to an AAF bin for a burden test. Each panel includes burden masks computed across 7 annotation categories. For binary traits, genotype counts among cases are displayed.

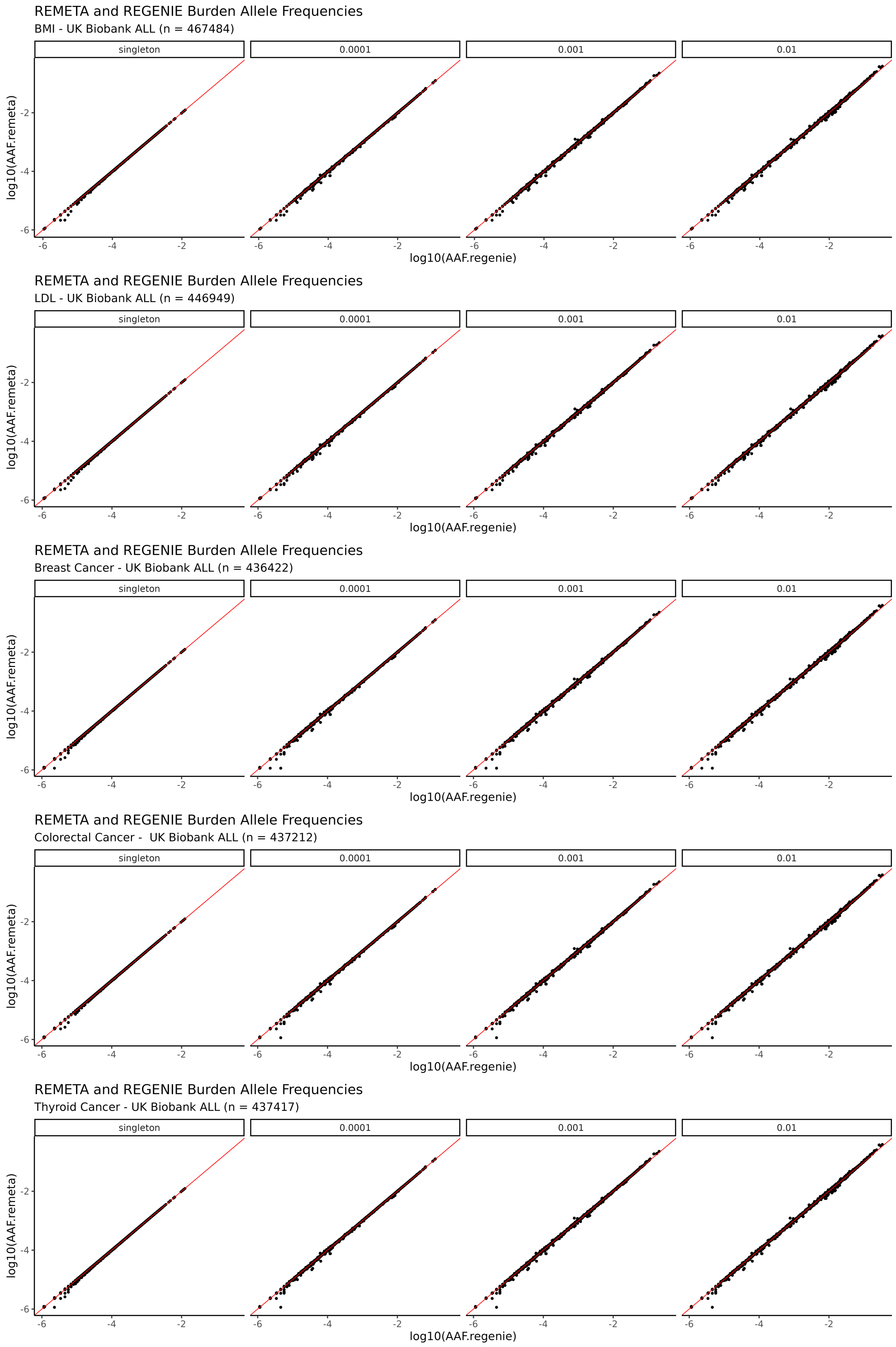

Supplementary Figure 10: **Scatterplot comparing allele frequencies of burden masks computed by REGENIE and REMETA**. Each row corresponds to a trait, and each column corresponds to an AAF bin for a burden test. Each panel includes burden masks computed across 7 annotation categories. The $x$-axis in each figure is the exact AAF computed in REGENIE, while the $y$-axis is estimated using the allele frequencies of the single variants in the test and the LD between them.

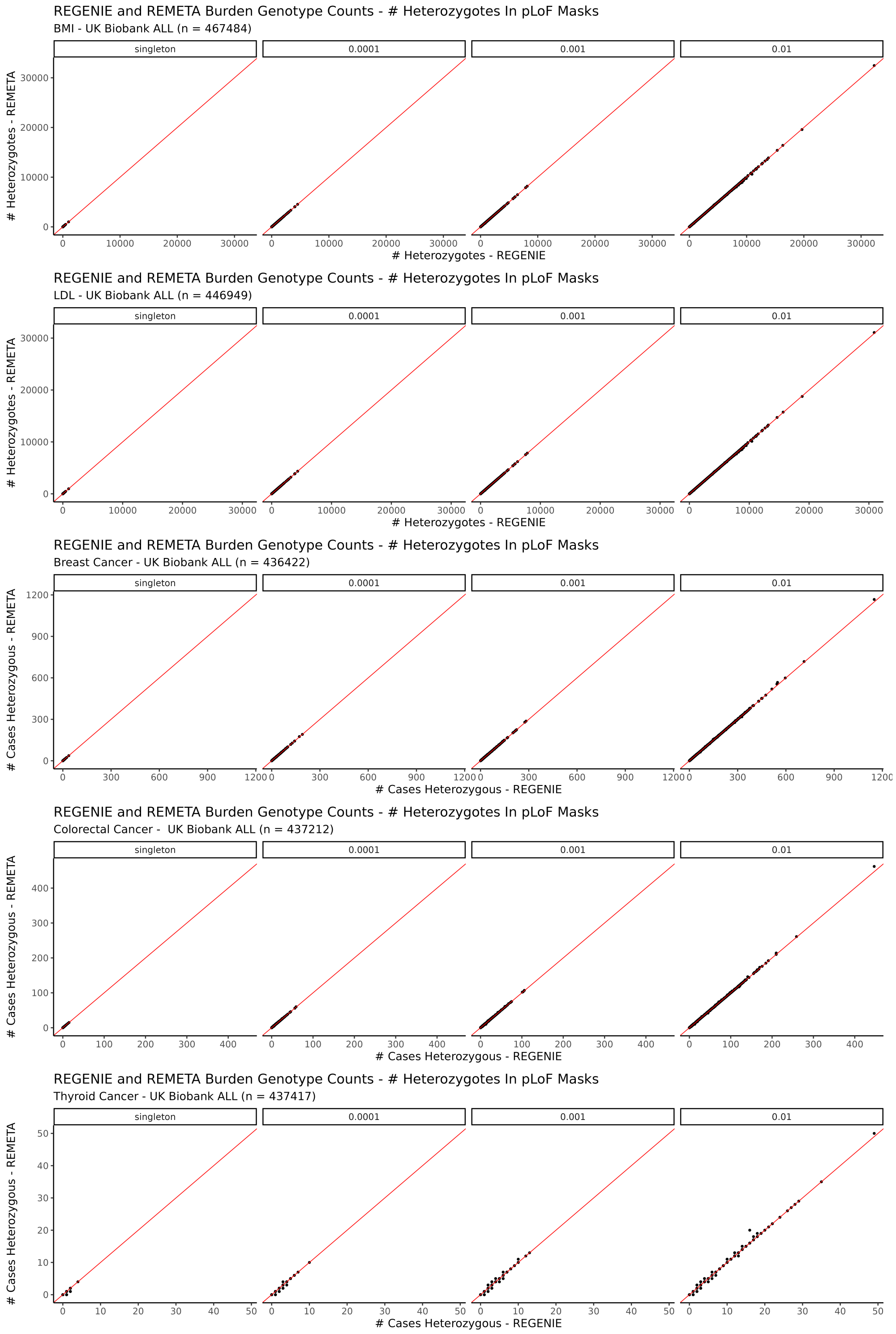

Supplementary Figure 11: **Scatterplot comparing genotype counts of burden masks computed by REGENIE and REMETA in pLoF masks.** Each row corresponds to a trait, and each column corresponds to an AAF bin for a burden test. Each panel includes only the pLoF mask for each gene. For quantitative traits, genotypes counts in the whole sample are displayed. For binary traits, genotype counts among cases are displayed.

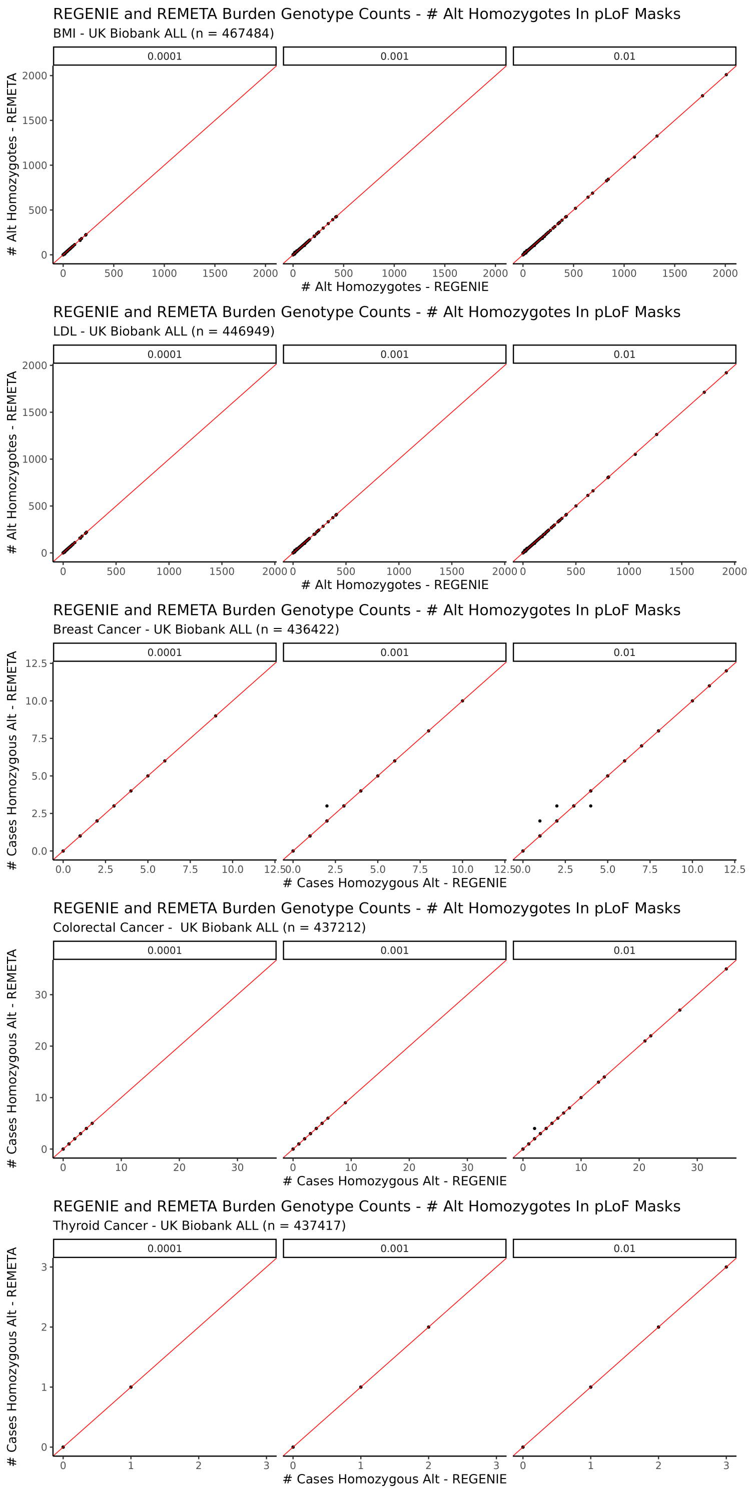

Supplementary Figure 12: **Scatterplot comparing genotype counts of burden masks computed by REGENIE and REMETA.** Each row corresponds to a trait, and each column corresponds to an AAF bin for a burden test. For quantitative traits, genotypes counts in the whole sample are displayed. Each panel includes only the pLoF mask for each gene. For binary traits, genotype counts among cases are displayed.

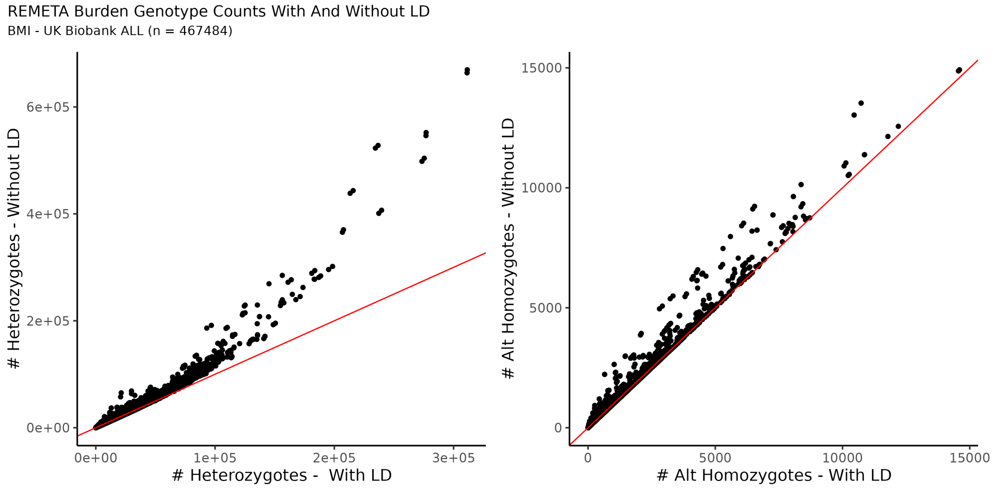

Supplementary Figure 13: **Scatterplot comparing genotype counts of burden masks computed REMETA to a naive estimate that ignores LD in BMI.** Estimates that ignore LD are equivalent to summing the genotype counts of the variants in mask.

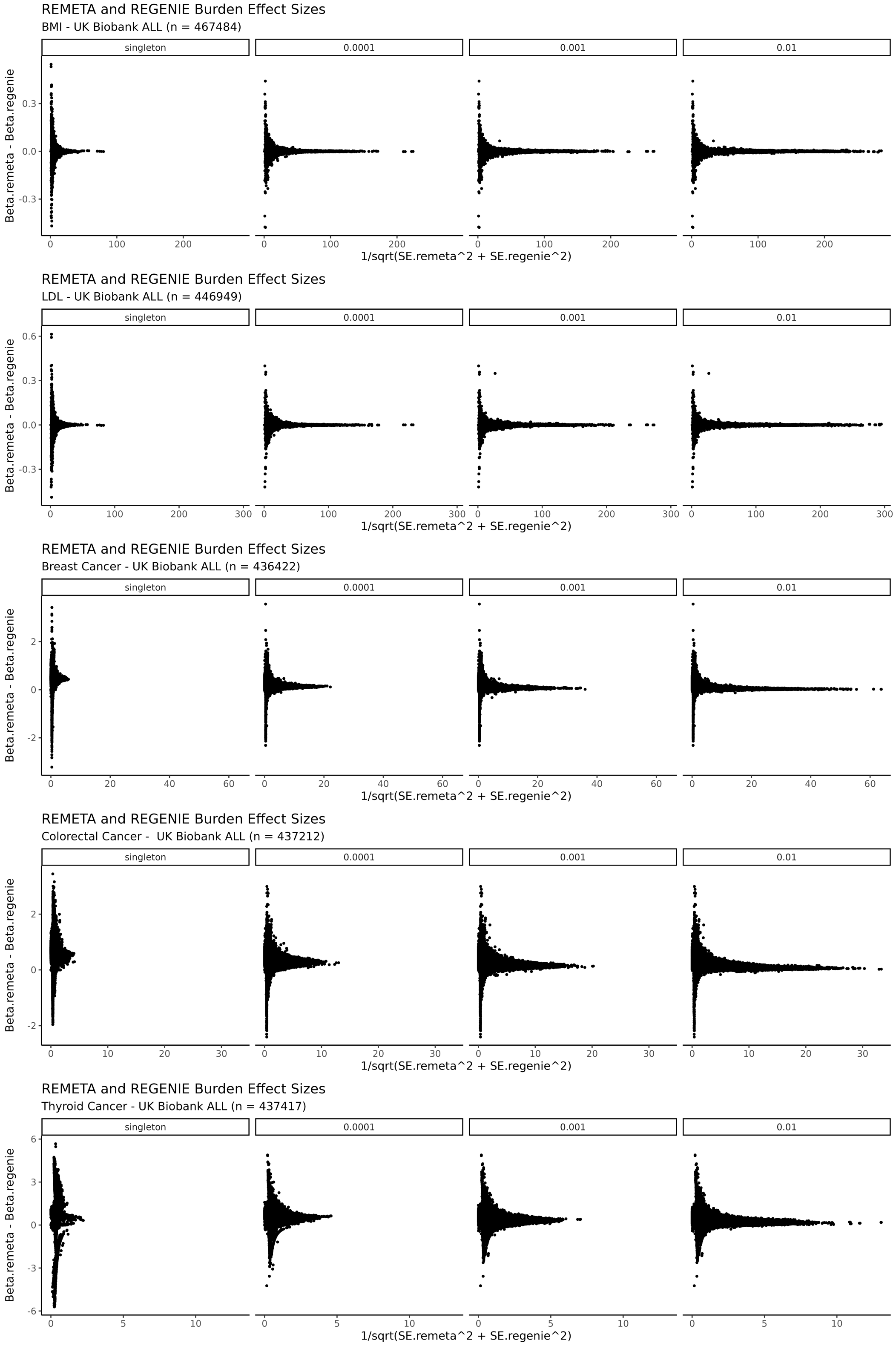

Supplementary Figure 14: **Comparison of effect size estimates from the sum test and the collapsing variant test.** Differences in effect size estimates between the sum test (Beta.remeta) and collapsing variant test (Beta.regenie). Note that the standard error of the difference $\beta.remeta - \beta.regenie$ is $\sqrt{\{SE^2_{remeta} + SE^2_{regene}\}}$. Each panel includes burden masks computed across 7 annotation categories.

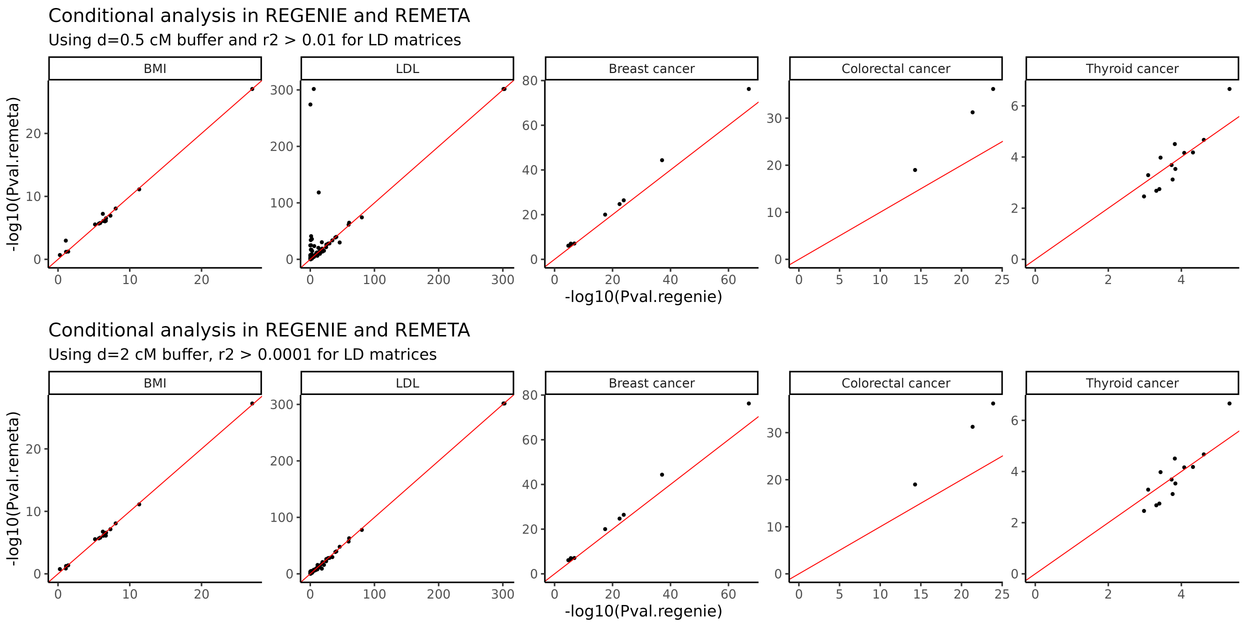

Supplementary Figure 15: **Scatterplot of conditional analysis p-values computed across two sets of parameters for LD matrix generation.** Top: Conditional analysis in REMETA is performed using LD matrices with a 0.5 cM buffer storing entries with $r^2 > 0.01$. Bottom: Conditional analysis in REMETA is performed using LD matrices with a 2 cM buffer storing entries with $r^2 > 0.0001$. Conditional analysis in REMETA is compared to conditional analysis in REGENIE. Except for LDL, most p-values computed by REMETA are similar between both parameter sets.

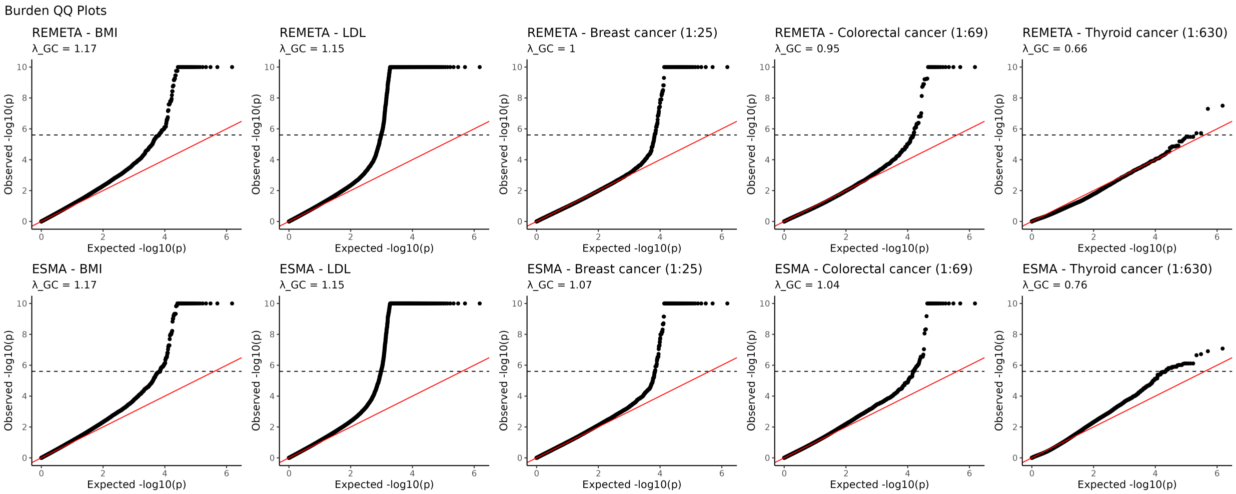

Supplementary Figure 16: **Burden test QQ plots from a meta-analysis of 3 subsets of UK Biobank.** Top: QQ plots of burden test $p$-values from meta-analysis with REMETA. Bottom: QQ plots of $p$-values from effect-size meta-analysis of burden tests. $\lambda_{GC}$: genomic control

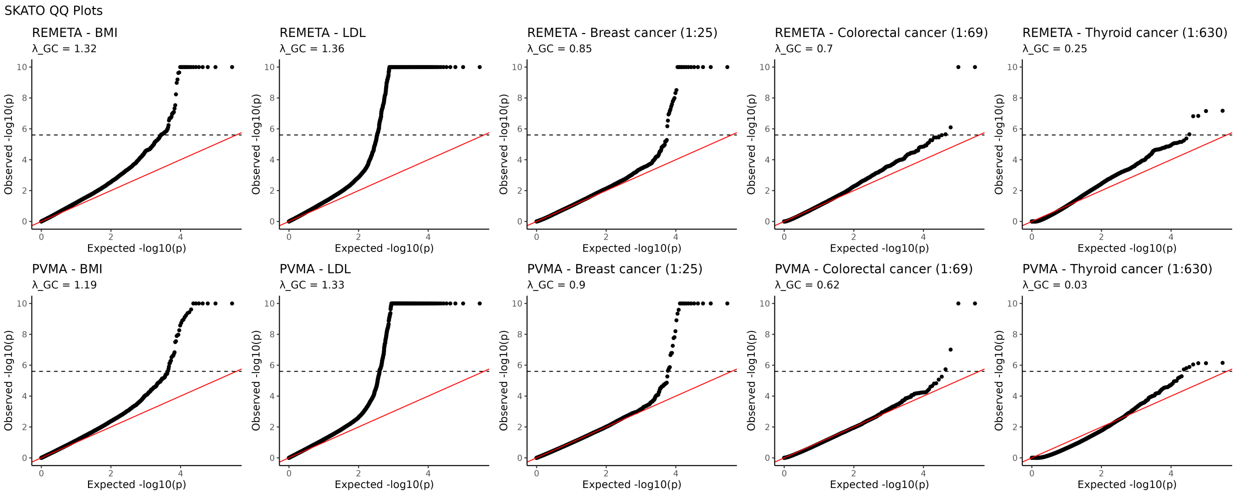

Supplementary Figure 17: **SKAT-O QQ plots from a meta-analysis of 3 subsets of UK Biobank.** Top: QQ plots of SKAT-O $p$-values from meta-analysis with REMETA. Bottom: QQ plots of $p$-values from p-value meta-analysis of SKAT-O. $\lambda_{GC}$: genomic control

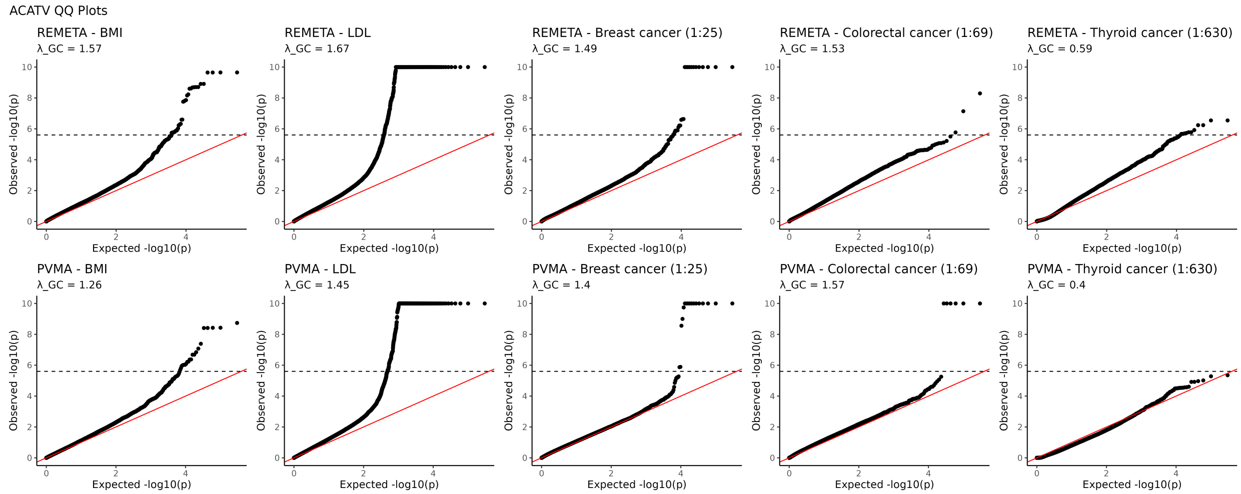

Supplementary Figure 18: **ACAT-V QQ plots from a meta-analysis of 3 subsets of UK Biobank.** Top: QQ plots of ACAT-V $p$-values from a meta-analysis with REMETA. Bottom: QQ plots of $p$-values from p-value meta-analysis of ACAT-V. $\lambda_{GC}$: genomic control

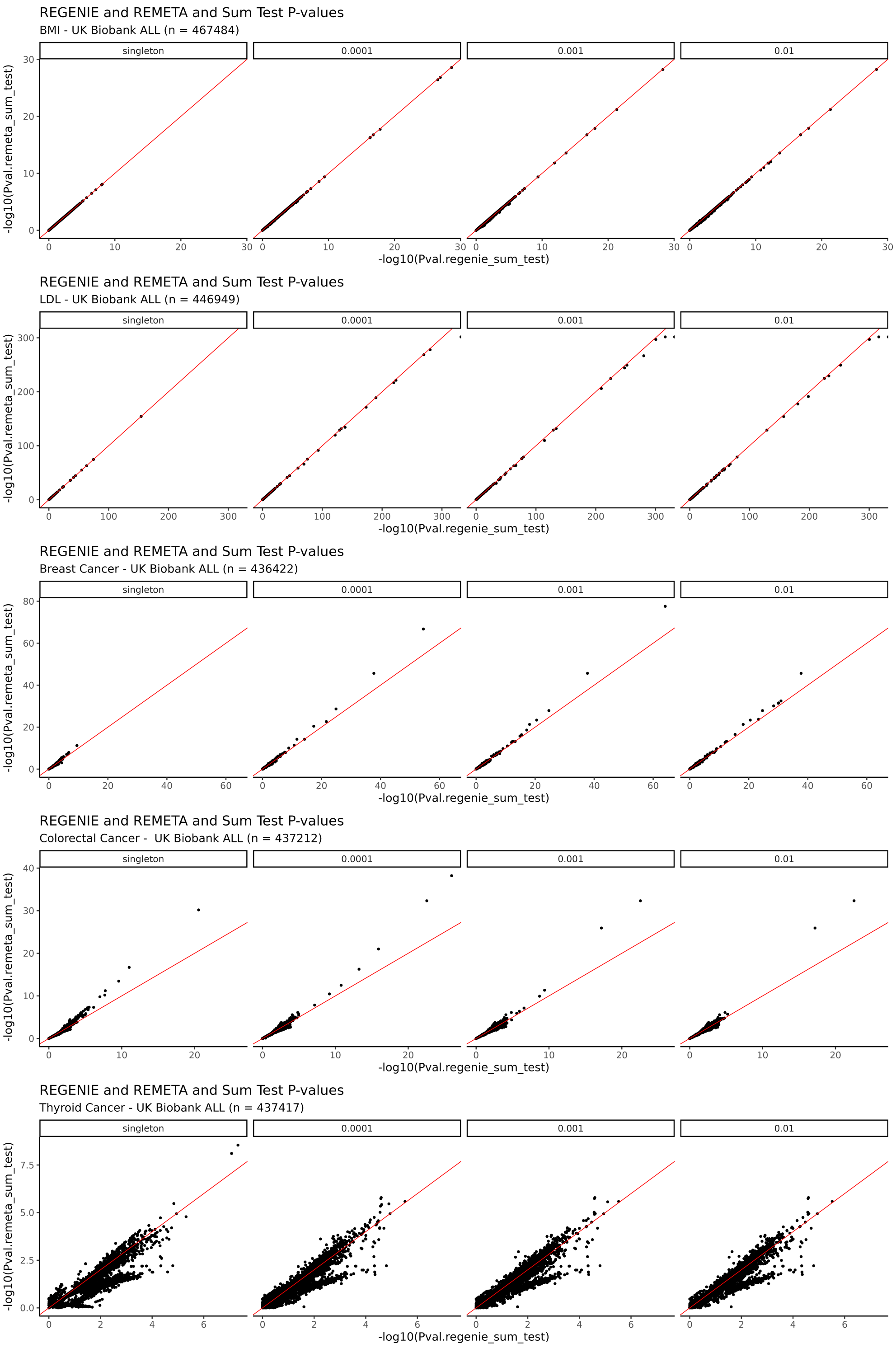

Supplementary Figure 19: **Scatterplot comparing $p$-value from the sum test computed using REGENIE and the sum test computed using REMETA.** Each row corresponds to a trait, and each column corresponds to an AAF bin for a burden test. Variants in an annotation category below an AAF threshold are grouped together for a test.

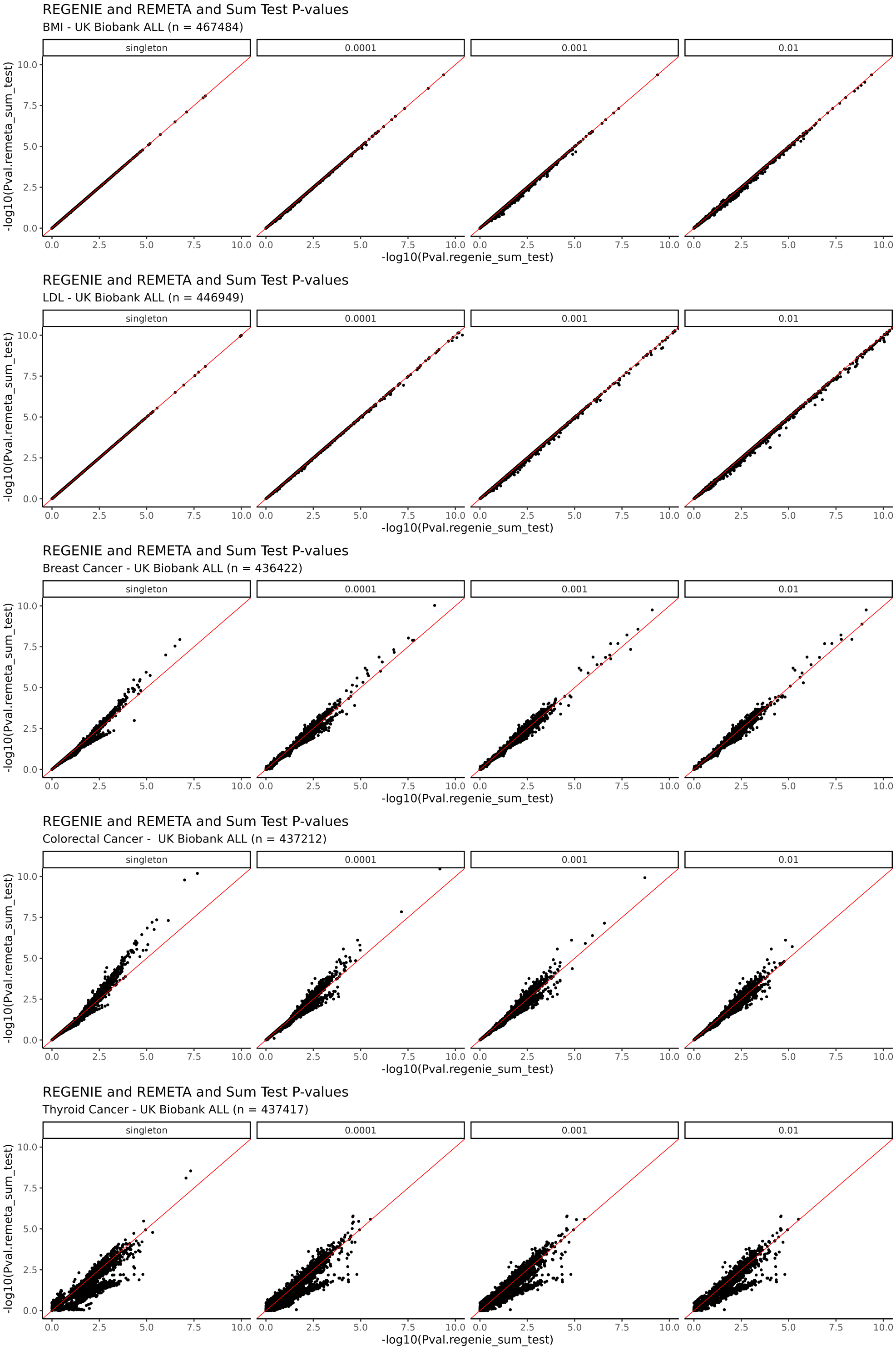

Supplementary Figure 20: **Zoomed-in scatterplot comparing $p$-value from the sum test computed using REGENIE and the sum test computed using REMETA.** Each row corresponds to a trait, and each column corresponds to an AAF bin for a burden test. Each panel includes burden masks computed across 7 annotation categories for each gene. Burden testing for the collapsing variant test and sum test was performed in REGENIE using the –build-mask max option.

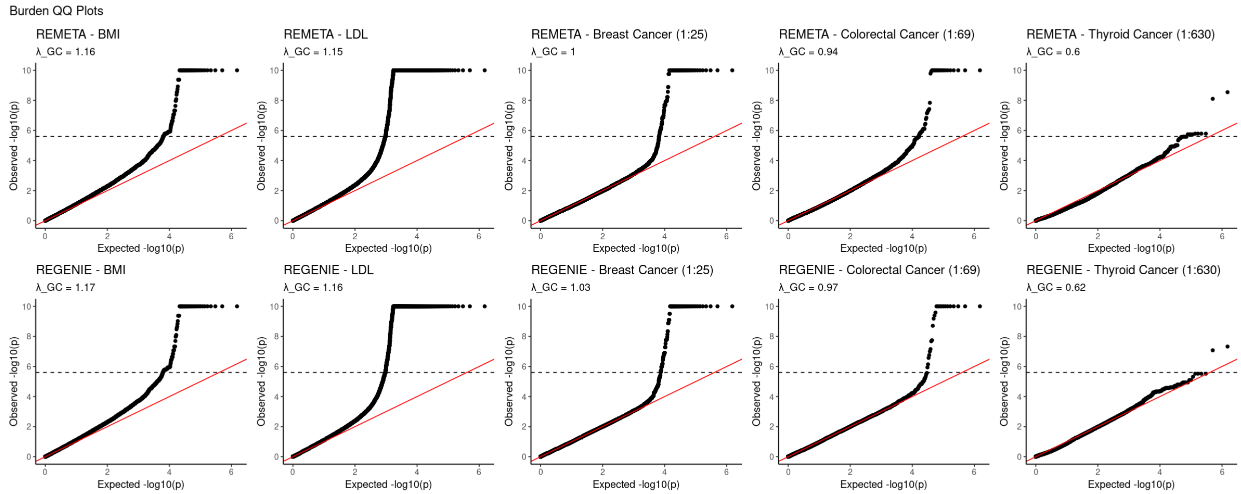

Supplementary Figure 21: **QQ plots from the sum test across 5 traits in UK Biobank ALL.** Top: QQ plots from the sum test computed with REMETA. Bottom: QQ plots from the sum test computed with REGENIE $\lambda_{GC}$: genomic control.

### 5 Supplementary Tables

| Trait | Burden | Burden-SPA | SKATO | SKATO-SPA |
|---|---|---|---|---|
| $\alpha = 1 \times 10^{-2}$ | | | | |
| QT | $0.952 \times 10^{-2}$ | - | $1.03 \times 10^{-2}$ | - |
| BT 1:1 | $0.963 \times 10^{-2}$ | $0.963 \times 10^{-2}$ | $0.998 \times 10^{-2}$ | $0.998 \times 10^{-2}$ |
| BT 1:9 | $0.940 \times 10^{-2}$ | $0.937 \times 10^{-2}$ | $1.13 \times 10^{-2}$ | $1.13 \times 10^{-2}$ |
| BT 1:49 | $0.919 \times 10^{-2}$ | $0.863 \times 10^{-2}$ | $1.57 \times 10^{-2}$ | $1.45 \times 10^{-2}$ |
| BT 1:99 | $0.948 \times 10^{-2}$ | $0.845 \times 10^{-2}$ | $1.88 \times 10^{-2}$ | $1.68 \times 10^{-2}$ |
| $\alpha = 1 \times 10^{-3}$ | | | | |
| QT | $0.995 \times 10^{-3}$ | - | $0.971 \times 10^{-3}$ | - |
| BT 1:1 | $0.981 \times 10^{-3}$ | $0.981 \times 10^{-3}$ | $0.946 \times 10^{-3}$ | $0.946 \times 10^{-3}$ |
| BT 1:9 | $1.03 \times 10^{-3}$ | $0.987 \times 10^{-3}$ | $1.34 \times 10^{-3}$ | $1.30 \times 10^{-3}$ |
| BT 1:49 | $1.24 \times 10^{-3}$ | $0.971 \times 10^{-3}$ | $2.83 \times 10^{-3}$ | $2.24 \times 10^{-3}$ |
| BT 1:99 | $1.37 \times 10^{-3}$ | $0.925 \times 10^{-3}$ | $3.94 \times 10^{-3}$ | $2.64 \times 10^{-3}$ |
| $\alpha = 1 \times 10^{-4}$ | | | | |
| QT | $0.910 \times 10^{-4}$ | - | $1.05 \times 10^{-4}$ | - |
| BT 1:1 | $0.857 \times 10^{-4}$ | $0.857 \times 10^{-4}$ | $0.927 \times 10^{-4}$ | $0.927 \times 10^{-4}$ |
| BT 1:9 | $1.17 \times 10^{-4}$ | $0.927 \times 10^{-4}$ | $1.64 \times 10^{-4}$ | $1.45 \times 10^{-4}$ |
| BT 1:49 | $1.85 \times 10^{-4}$ | $1.07 \times 10^{-4}$ | $5.41 \times 10^{-4}$ | $3.17 \times 10^{-4}$ |
| BT 1:99 | $2.55 \times 10^{-4}$ | $1.07 \times 10^{-2}$ | $9.39 \times 10^{-4}$ | $4.83 \times 10^{-4}$ |

Supplementary Table 1: **Evaluation of type 1 error rates for burden tests and SKAT-O.** Each cell represents the empirical type 1 error averaged across 10 simulation replicates. Null traits were simulated across three equally sized subsets of UK Biobank and meta-analyzed with REMETA. Gene-sets were constructed across three annotations categories—pLoFs, all missense variants, and synonymous variants—using an allele frequency cutoff of 1%. Overall, 57,165 gene-based tests were performed for each trait.
